# Interpretable competing-risk prediction of hepatocellular carcinoma in cirrhosis from region-wide routine health data

**DOI:** 10.64898/2026.11.19.26368516

**Authors:** Isidoro Gutiérrez-Álvarez, Ángela Rojas, Luis Calzadilla-Bertot, Joaquín Dopazo, Manuel Romero-Gómez, Carlos Loucera

## Abstract

**Background:** Short-term prediction of hepatocellular carcinoma (HCC) in cirrhosis is complicated by the competing risk of death from liver disease. We aimed to develop an interpretable model on region-wide routine data that predicts HCC while separating competing mortality, and to compare it with established risk scores.

**Methods:** From a region-wide cohort of adults with cirrhosis in the Andalusian Population Health Database, we trained the model on 17,177 patients with a prediction date set 12 months before their outcome, between 2020 and 2023. From one year of electronic health records before that date (demographics, liver and chronic disease codes, and eight routine laboratory tests), an Explainable Boosting Machine without interaction terms classified the following 12 months as HCC, death or liver transplantation without HCC, or event-free. We evaluated it in 20 repeats of 5-fold cross-validation against a binary model, a model with pair interactions and a model using the full health record, all on the same folds. We then applied the cross-validated models, without retraining, at the first routine encounter at which aMAP or the Toronto HCC Risk Index could be computed, and compared them with the scores in patients followed forward under competing risks, with and without a 6-month washout and with each province left out of training in turn.

**Findings:** 771 patients (4.5%) developed HCC within 12 months and 3,314 (19.3%) died or underwent liver transplantation without HCC (3,061 deaths, 253 transplants). In cross-validation, the area under the receiver operating characteristic curve (AUROC) for HCC was 0.734 (SD 0.018). A binary formulation, pairwise interactions and the full health record (more than 3,000 features) did not improve it; the full record lowered it by 0.024. At routine encounters the model discriminated 12-month HCC better than aMAP (time-dependent AUC 0.790 vs 0.688; difference +0.102, 95% CI +0.071 to +0.132) and the Toronto HCC Risk Index (0.761 vs 0.703; +0.057, +0.032 to +0.081), and remained better after excluding HCC diagnosed within 6 months (concordance difference +0.055 and +0.040) and when each of the eight provinces was left out of training. At equal group size, its high-risk group held as much HCC as the scores’ high-risk categories, with 20–28% fewer competing deaths. Its shape functions separated cancer-specific signal (AFP rising from 2 to 20 µg/L, platelets below 150 ×10^9^/L) from liver failure (bilirubin above 51 µmol/L, albumin below 35 g/L); a binary model showed only the first.

**Interpretation:** An interpretable three-class model on routine data outperformed established scores for short-term HCC risk in cirrhosis, while showing whether each abnormal value points to cancer or to competing death.

**Funding:** This work was supported by grants PID2023_152380OB and PRTR-C17.I1 from the Spanish Ministry of Science and Innovation; grant PMP24/00005 from the Instituto de Salud Carlos III (ISCIII); grant IE19_259 FPS from Consejería de Salud y Consumo, Junta de Andalucía; and Horizon Europe (HORIZON), Marie Skłodowska-Curie Actions Doctoral Networks (101226456-MLCARE). The funders had no role in study design, data collection, data analysis, data interpretation or writing of the report.

**Research in context:** *Evidence before this study:* We searched PubMed to October 2026 for HCC risk prediction in cirrhosis with machine learning or risk scores. Machine-learning models usually outperform aMAP and the Toronto HCC Risk Index (THRI) in development data. They are mostly specific to one etiology (HBV or HCV), built on inputs not routinely available (elastography), black boxes explained post hoc, and evaluated without competing risks. A meta-analysis of models for non-viral liver disease found only one developed with a competing-risk approach, and rated all 14 included studies at high risk of bias (1). In prospective French cohorts analysed under competing risks, machine learning did not outperform aMAP (2).

*Added value of this study:* In a region-wide population of 17,177 adults with cirrhosis of all etiologies, we developed an interpretable three-class model on routine records: HCC, death or transplant, or event-free. Under competing risks, at the first encounter where each score could be computed, it ranked HCC better than aMAP and THRI, including after a 6-month washout. At the same group size it flagged as much HCC with 20–28% fewer patients who died first. The advantage held when each province was left out of training. Its shape functions are the whole model, not a post-hoc approximation, and separate markers of cancer from markers of liver failure. A binary model on the same data ranks HCC as well but folds the death signal into its single curve, as we show analytically.

*Implications of all the available evidence:* Surveillance programmes can rank patients from data they already hold, with a model whose every contribution can be inspected. Modelling death as an outcome keeps patients unlikely to benefit off the high-risk list. External validation in other health systems, with local recalibration, is the next step.

## Introduction

Hepatocellular carcinoma (HCC) is a major cause of cancer-related mortality (3) and usually develops in the context of chronic liver disease, most often cirrhosis (4). Surveillance seeks to detect HCC at a treatable stage, but patients with cirrhosis do not share the same near-term risk: some develop HCC within months, whereas others face a more immediate risk of hepatic decompensation or death (5). Short-term prediction in cirrhosis is therefore not simply a cancer-versus-non-cancer problem; it requires distinguishing HCC risk from competing mortality risk within the same clinical time window (6).

Recent clinical prediction and machine-learning studies show that routinely available data can support HCC risk stratification across diverse clinical settings. Conventional models, including aMAP (7), the Toronto HCC Risk Index (THRI) (8) and the Kanwal cirrhosis score (9), use routinely measured clinical and laboratory variables to stratify HCC risk in chronic liver disease and cirrhosis, while deep-learning models built on longitudinal electronic health records (EHRs) have identified patients with cirrhosis at high HCC risk (10). Machine-learning models often outperform these scores in development data, but are typically binary, evaluated without competing risks, or built on inputs not routinely available; in prospective French cohorts analysed under competing risks, machine-learning models did not outperform aMAP (2).

Population-level frameworks such as PRE-Screen-HCC (11) extend this approach upstream, using large cohorts to identify individuals at increased HCC risk before chronic liver disease or cirrhosis is consistently recognised. This addresses an important screening gap because cirrhosis is often unrecognised, or recognised only shortly, before HCC diagnosis (12), and some HCC arises without cirrhosis (13). However, patients with established cirrhosis are already candidates for surveillance, unless competing mortality is high or curative treatment is not feasible (4), and present a different decision problem: their short-term HCC risk must be interpreted alongside competing risks of decompensation and death, since binary HCC-versus-non-HCC modelling may obscure clinically distinct trajectories by grouping patients who die without HCC with those who remain alive and event-free (5,6).

Region-wide real-world data allow this problem to be studied at the scale required for health-system deployment. The Andalusian Population Health Database (BPS) is well suited to this task because it integrates longitudinal EHRs from more than 15 million patients in the Andalusian public health system (14), including diagnoses, laboratory results, imaging, prescriptions, hospital activity, and mortality-related information (15). Its secondary use is enabled through PAGEM, a secure processing environment embedded within the healthcare network, where pseudonymised data are analysed under GDPR-compliant and EHDS-aligned governance, and only non-identifiable outputs are released (14). This framework supports longitudinal studies of incidence (16), treatment use and outcomes (17), survival (18), drug repurposing (19–21), synthetic EHR generation (22), and predictive modelling across the regional public health system (23).

In such high-stakes settings, interpretability is central to clinical translation (24). A short-term HCC prediction model should not only estimate risk, but also show whether abnormal biomarkers support cancer risk, hepatic deterioration, or event-free survival. Explainable Boosting Machines are well suited to this task because they learn feature-specific response functions that can be inspected directly (25). This allows clinicians to assess whether the model recovers plausible biomarker patterns, whether abnormal values reflect HCC risk or competing mortality, and whether clinically familiar thresholds emerge directly from real-world data rather than being imposed during model development.

To this end, we developed an interpretable machine-learning model for 12-month HCC prediction in patients with cirrhosis using routine public health data from Andalusia. We framed the task as a three-class prediction problem: incident HCC, death or liver transplantation without HCC, and event-free survival. Using an Explainable Boosting Machine trained on a curated clinical feature set, we evaluated discrimination, precision-recall performance and class-specific feature-response profiles in repeated stratified cross-validation. We compared the model, on the same cross-validation folds, with a binary HCC model, a model with pairwise interaction terms, and a model using the full health record (3,000+ features), with variance-corrected paired tests and Bayesian equivalence testing (26,27). We then compared it with aMAP and THRI at the first routine encounter at which each score could be computed, under competing risks, and recalibrated it to absolute risk at routine laboratory visits. Because HCC risk and its predictors differ by sex and etiology, we report performance within these groups. Crucially, we evaluated the clinical plausibility of learned biomarker shape functions: standard binary formulations group deaths without HCC with event-free survivors, so a low predicted risk cannot distinguish a healthy patient from one about to die of liver failure, whereas a three-class model shows the cancer and the competing-death signals separately. Finally, to support reproducibility and external validation, we transformed the data pipeline to OMOP-compatible structures and provide code for feature extraction, preprocessing, model training, and evaluation, enabling other institutions to reproduce the feature definitions and port the model to harmonised EHR environments.

## Methods

### Data source and ethics

Real-world longitudinal data were extracted from the Population Health Database (15), known in Spanish as Base Poblacional de Salud (BPS), which aggregates electronic health records (EHRs) from all users within the Andalusian public health system. The study was approved by the Ethics Committee for the Coordination of Biomedical Research in Andalusia (CCEIBA; Acta 10/24). All data were transferred from the BPS systems (pseudonymised in origin) to the PAGEM trusted research environment (14), where all analyses were performed.

### Study design

We designed this retrospective observational cohort study using routinely collected healthcare data from the BPS: one cohort, analysed in two designs that differ only in their time zero (Fig. 1 a). In the development design, the model was trained and internally validated by repeated cross-validation on prediction dates anchored on each patient’s outcome: a sampling of time zero that is efficient for learning how the predictors relate to the three outcomes, but that does not reproduce the case mix met in practice. In the evaluation design, the same cross-validated models, without retraining, were applied at routine encounters, dates that do not depend on the outcome, and patients were followed forward under competing risks. The evaluation estimates, not the cross-validated ones, describe performance at the point of care; they underlie the comparison with aMAP and THRI and the calibration of the deployment model. Throughout, time zero is the date from which features look back and outcomes look forward: the prediction date in development, the landmark or laboratory visit in evaluation.

**Fig. 1.**
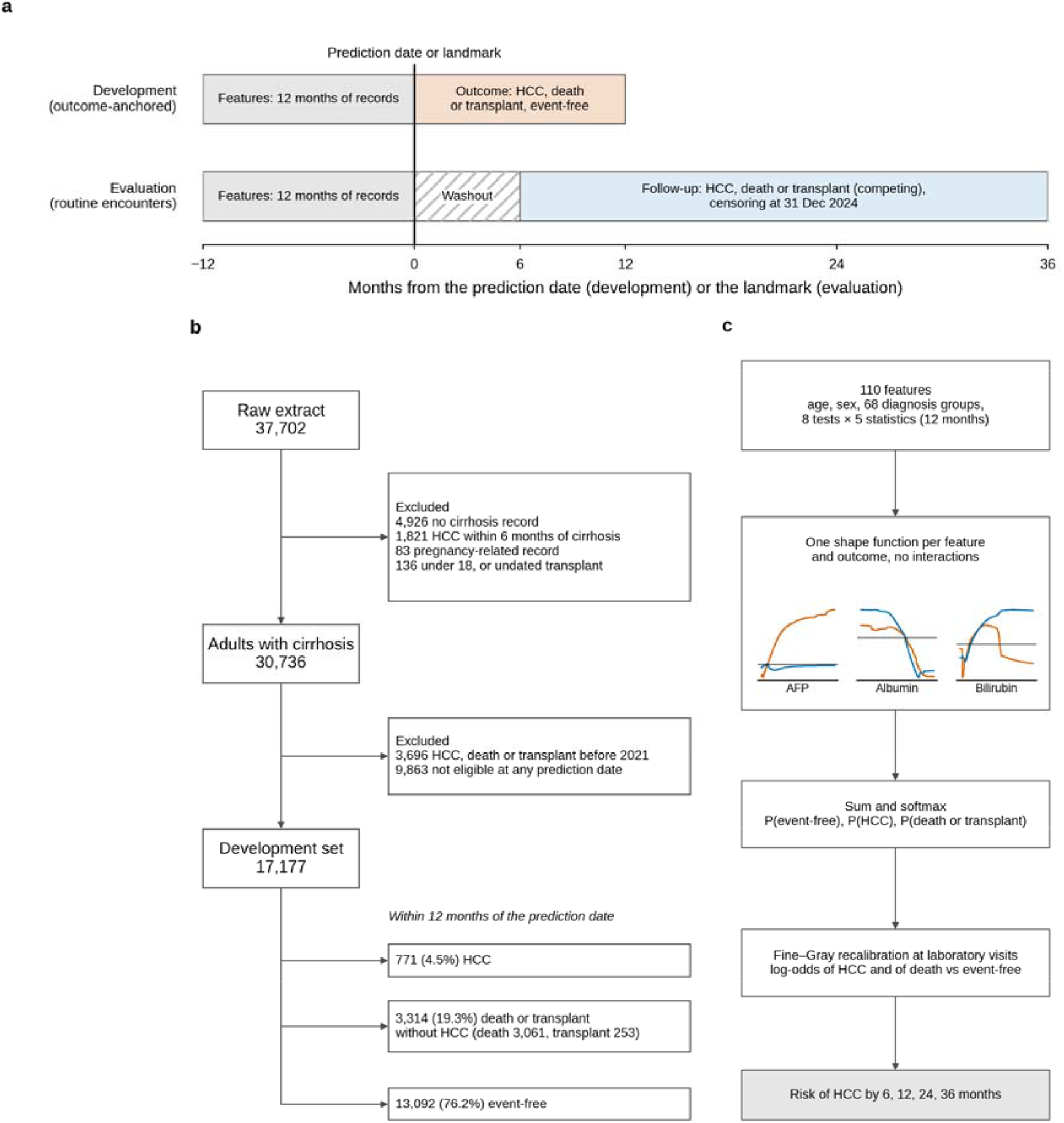
Study design, cohort and model. **a**, The two designs on one time axis. Development, outcome-anchored (top): for each patient, the prediction date lies 12 months before the index date (the date of HCC, of death or liver transplantation without HCC, or a date drawn from the cases’ index dates for event-free patients); features come from the 12 months of records before it, and the outcome is the first event in the following 12 months. Evaluation at routine encounters (bottom), used for the comparison with aMAP and THRI: each score’s landmark is the first date, from 2 January 2020, at least 6 months after the cirrhosis record, on which all of its laboratory components were measured; the model’s features are rebuilt from the 12 months before the landmark, and follow-up runs to HCC, death or liver transplantation (competing events) or to 31 December 2024. The 6-month washout (hatched) applies only to the sensitivity analysis: patients with an event or censoring in it are excluded, and time restarts at its end. **b**, Flow of patients to the development set (the landmark sets, largely the same patients, are in Supplementary *Table 1*). Not eligible at any prediction date: cirrhosis recorded less than 6 months earlier, or a previous cancer, TIPS or liver transplant at every candidate date from 2020 to 2023. Deaths and liver transplants without HCC form one outcome class, shown here separately. **c**, The deployment model. Each of the 110 features, derived from age, sex, coded diagnoses and eight laboratory tests, enters through one shape function per outcome, with no interaction terms. Sparklines show the reference model’s contrasts for AFP, albumin and bilirubin: HCC (vermilion) and death or transplant (blue), each against event-free. The contributions add up and pass through a softmax to three class probabilities. A Fine–Gray model, fitted at routine laboratory visits, maps the two log-odds against event-free to the absolute risk of HCC by 6 to 36 months (Methods).

### Cohort definition

We used records from 1 January 2019 to 31 December 2024, together with chronic-disease diagnoses of any date from the BPS chronic-condition registry. Adults with cirrhosis were identified by ICD-10-CM codes K70.3, alcoholic cirrhosis; K71.7, toxic liver disease with fibrosis and cirrhosis; or K74, fibrosis and cirrhosis of the liver (and their ICD-9-CM equivalents, translated with the CMS General Equivalence Mappings). We defined incident HCC as a diagnosis coded as ICD-10-CM C22.0. Of 37,702 patients, we excluded 4,926 without a cirrhosis code, 1,821 whose HCC was diagnosed within 180 days of the cirrhosis record, 83 with a pregnancy-related code and 136 who were under 18 at the cirrhosis record or had an undated liver-transplant complication, leaving 30,736 (Fig. 1 b). Both designs applied the same eligibility rule at time zero: at least 180 days past the first cirrhosis record, alive, free of HCC, and without a previous cancer, a transjugular intrahepatic portosystemic shunt (TIPS) or a liver transplant. No sample size calculation was needed: both designs draw on the whole population with a recorded cirrhosis diagnosis in the Andalusian public health system who met the eligibility criteria, not a sample of it. The precision of each estimate is given by its spread over the repeated cross-validation or by its confidence interval.

### Development design: outcome-anchored prediction dates

Each patient received an index date: the date of HCC for patients who developed it, the date of death or liver transplantation for those who died or were transplanted without HCC, and for the rest a date drawn at random from the cases’ index dates. Index dates ran from 2021 to 2024. The prediction date lay 12 months before the index date, and features came from the 12 months before it, so no record from the prediction window was used (Fig. 1 a). Of the 30,736, 3,696 had HCC, died or were transplanted before 2021, and 9,863 were not eligible at any prediction date (cirrhosis recorded less than 6 months before it, or a previous cancer, TIPS or liver transplant), leaving 17,177 patients with an eligible prediction date (the development set; Fig. 1 b), each with a 12-month three-class outcome.

### Evaluation design: landmarks at routine encounters

For each score, the landmark was the first date, from 2 January 2020, on which all of the score’s laboratory components were measured with the patient eligible: 14,928 patients had an aMAP landmark and 18,848 a THRI landmark (the landmark sets; Supplementary Table 1). They were followed to HCC, death or liver transplantation, or to 31 December 2024 (Comparison with established risk scores). The recalibration of the deployment model used routine laboratory visits, the first eligible visit per patient in each 6-month window from 2020 to 2024 (82,134 visits; Deployment model and absolute-risk recalibration). The landmark sets are largely the same patients as the development set (13,048 of the 14,928 with an aMAP landmark and 16,013 of the 18,848 with a THRI landmark also had a prediction date), seen at a different time zero. The evaluation is therefore internal, in the same population and period, not a validation in new patients; each patient was scored by a cross-validation model that had not been trained on them.

### Predictor variables

We extracted demographic, chronic condition, and laboratory variables from the electronic health record. All variables were retrieved from the 12 months before time zero.

The sociodemographic variables were date of birth and sex.

We used chronic disease records curated by BPS from ICD-10 codes and free-text information documented in the EHR. ICD-9-CM diagnoses were translated to ICD-10-CM before grouping. Supplementary Table 7 lists the features.

The laboratory variables include eight key liver function and haematological biomarkers specifically selected for their clinical relevance to hepatocellular carcinoma risk assessment: platelet count (OMOP concept_id: 3024929), platelet mean volume (OMOP concept_id: 3001123), alanine aminotransferase (ALT, OMOP concept_id: 3006923), total bilirubin (OMOP concept_id: 3006140), prothrombin time (PT, OMOP concept_id: 3034426), aspartate aminotransferase (AST, OMOP concept_id: 3013721), albumin (OMOP concept_id: 3024561) and alpha-1-fetoprotein (OMOP concept_id: 3039783).

The model therefore needs only data already held in the health record: age, sex, the coded chronic diagnoses and eight routine laboratory tests. From these, the pipeline derives 110 features automatically: age, sex, 68 diagnosis-group flags, and five summary statistics (mean, median, SD, maximum, minimum) of each of the eight tests. We restricted the feature set to variables with prior use in HCC risk models. Nevertheless, we report a complementary model using the full set of variables extracted from the electronic health record (see Comparison of model variants).

### Feature engineering

We converted the recorded variables into model features using only information recorded in the 12 months before time zero. This ensured that no data from after time zero (the prediction window in development, the follow-up in evaluation) contributed to feature construction. Predictors are routine laboratory results and coded diagnoses, extracted automatically, so no subjective interpretation was involved.

We used the date of birth to calculate age in days at time zero: the prediction date in development, and the landmark, the laboratory visit or the date of use in evaluation and deployment. Sex was encoded as a binary variable (1 indicating male).

For diagnostic variables, ICD-10 codes were grouped into blocks, except for conditions in block K70–K77 (Diseases of the Liver), which were further expanded to the category level, i.e., the first three characters of each code. This approach reduced dimensionality while preserving clinically interpretable diagnostic categories. For each grouped ICD-10 category, we created a binary indicator reflecting whether the patient had at least one corresponding diagnosis recorded before time zero.

For laboratory variables, we summarised all available measurements recorded in the 12 months before time zero by the mean, median, standard deviation, minimum, and maximum value. A missing value took the training median.

### Outcome definition

We modelled the prediction task as a three-class outcome to account for the competing risk of death. In development, each patient was assigned to one of three mutually exclusive outcome groups over the 12 months after the prediction date: HCC; death or liver transplantation without HCC; event-free. In evaluation, the same outcomes were analysed as times from time zero to the first of HCC or death or liver transplantation without HCC (competing events), censored at 31 December 2024 and followed up to 36 months. A liver transplant without previous HCC ends the natural history of the liver disease as death does, so it is counted with death. Outcomes were derived automatically from coded records, independently of the predictors, so no subjective assessment or blinding of assessors was involved.

### Model development

We trained the main model as a three-class classifier using the structured feature set derived from the EHR. We used an Explainable Boosting Machine (EBM) (25) as the primary modelling approach. We used the default EBM hyperparameters except that pairwise interaction terms were disabled (interactions = 0), so that the model is a sum of one shape function per feature and its global explanation is complete. (For three classes, the interpret library does not display pairwise terms, so a model with them would be only partly visible.)

We addressed outcome imbalance using class weights estimated within each training subset. For each cross-validation iteration, we calculated class weights according to the training-specific outcome prevalence, so that less frequent classes received larger weights during model fitting.

We implemented preprocessing and model training within a single reproducible pipeline. The pipeline applied standard scaling to age; sex and diagnostic indicators entered as binary flags. For laboratory measurements, we applied robust scaling based on the interquartile range to reduce the influence of extreme values, then median imputation. Supplementary Table 8 lists every preprocessing step and hyperparameter with its value.

We evaluated model performance using 20 repeats of 5-fold stratified cross-validation, yielding 100 train-test evaluations. Folds were stratified by the three-class outcome and seeded, so every model below was trained and tested on the same 100 folds of the same patients. Within each iteration, we held out one fold as the test set and split the remaining data internally, reserving 20% as a validation subset for threshold selection. We trained the model on the training subset, selected the operating threshold on the validation subset, and evaluated performance on the held-out test fold. The preprocessing parameters learned on the training subset were applied unchanged to the validation and test subsets.

The model produced class-specific predicted probabilities for HCC, death or transplantation without HCC, and event-free survival. For the HCC class, we selected the operating threshold to target a false-positive rate of 10% in the validation subset. We then applied the selected threshold to the corresponding test fold.

We assessed discrimination for HCC prediction using the area under the receiver operating characteristic curve (AUROC) and the area under the precision-recall curve (AUPRC), treating HCC as the target class against the remaining outcome classes. Because every patient’s 12-month outcome class is known, these are ordinary, time-fixed AUROC and AUPRC, without censoring weights; we keep the term AUROC for them, and AUC( ) for the time-dependent AUC of the comparison with the scores. We also evaluated threshold-dependent performance using sensitivity, specificity, positive predictive value, negative predictive value, F1 score, balanced accuracy, Matthews correlation coefficient, and the likelihood ratios. We calculated all threshold-dependent metrics in each test fold using the threshold selected in the corresponding validation subset. We reported the mean and standard deviation of each metric across the 100 test evaluations.

### Model formulation and interpretation of shape functions

For patient features x = (x_1_ , …, x_p_) the model computes, for each class k (0 event-free, 1 HCC, 2 death or transplantation),

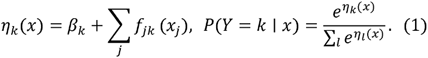

Each *f_jk_* is a step function over the bins of feature ,. A shift added to all three classes’ functions leaves the probabilities unchanged, so only contrasts between classes are identified. We therefore show, for each feature, 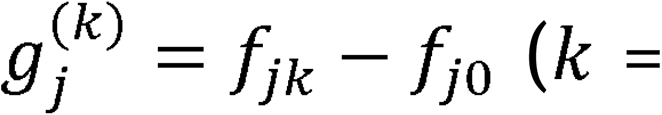 HCC, death/transplant): the change in the log-odds of that outcome against staying event-free, so that

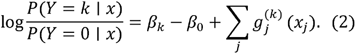

Each curve is centred to mean zero over the training patients and summarised as mean ± SD over the 100 cross-validation models. A laboratory test enters through five statistics of the 12-month window (mean, median, SD, maximum, minimum); we plot the median’s function and add the five when ranking feature groups.

Training used balanced class weights ,w_k_ = n/(3n_k_). For a well-specified model these shift only the intercepts,

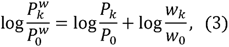

and leave the shape functions unchanged. The raw probabilities therefore rank patients but overstate risk: 17-fold odds for HCC (log17 - 2.83).

The binary model’s target pools death with event-free. Its log-odds are log[P_HCC_/ (P_EF_ + P_D_)] - 1_H_ — log(1 + e^LD^), with *L*_k_ the log-odds of class *k* against event-free, so, approximately, its shape function *h_j_* satisfies

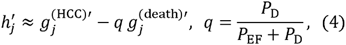

with q the probability of death among patients without HCC (Supplementary Methods).

### Comparison of model variants

We compared the reference model (three classes, curated features, no interactions) with three variants, each changing one element: a binary model (HCC against the rest, on the same patients and folds), the same model with interpret’s default pairwise interactions, and a model using the full health record. The full record comprised every diagnosis (ICD-10-CM truncated to three characters, registry and other sources separately), procedure (ICD-10-PCS, four characters), laboratory test (five summary statistics and the slope), active drug ingredient and visit type: more than 3,000 features in all, without interactions.

To compare model performance while accounting for the dependence between cross-validation folds, we used Nadeau and Bengio’s corrected paired t-test (26); this variance-corrected approach addresses the issue of non-independent samples arising from models being evaluated on the same data partitions. For multiple comparisons, we applied Bonferroni correction over the three comparisons to control the family-wise error rate.

In addition to the frequentist approach, we conducted Bayesian analysis to compute the posterior distribution of performance differences between models. This allowed us to calculate the probability that one model is better than another, as well as the probability that models are practically equivalent within a region of practical equivalence (ROPE) of ±0.01 (27).

### Comparison with established risk scores

We compared the model with aMAP (7) and THRI (8), computed from routine BPS data with the published formulas and categories (aMAP: low < 50 < medium < 60 < high; THRI: low < 120 < intermediate < 240 < high). THRI’s etiology came from the chronic-condition codes recorded by the landmark; with more than one etiology the highest-scoring one was used (Supplementary Table 9). Sustained virological response after hepatitis C cannot be identified in these data, so hepatitis C was scored as viraemic (main analysis) and as cured (Supplementary Table 5), which bounds each patient’s score. The Kanwal score was not compared: body mass index is not recorded and smoking is recorded too sparsely.

Each score was evaluated at its own landmark: the first date, from 2 January 2020 and at least 180 days after the cirrhosis record, on which all of its laboratory components were measured, with the patient eligible as above (Fig. 1 a; Supplementary Table 1). At the landmark the model’s features were rebuilt from the 12 months before it, and each patient was predicted by the cross-validation model that had not seen them (patients outside the training set by a fold assigned at random per repeat), giving one out-of-fold prediction per patient in each of 20 repeats, without retraining. Follow-up ran to HCC, death or liver transplantation (competing events), or 31 December 2024. All comparisons were paired on the same patients:

1. discrimination: the cumulative/dynamic time-dependent AUC for HCC at = 6, 12, 24 and 36 months, AUC( ), estimated with inverse probability of censoring weights (IPCW), with patients who died or were transplanted by counted as controls (28). Unlike the AUROC above, AUC( ) handles censored follow-up and the competing events. We report it with 95% CIs from 200 paired bootstrap resamples, and the difference in cause-specific concordance to 36 months (29);
2. sensitivity for HCC when each marker flags the same fraction of patients;
3. stratification: Aalen–Johansen cumulative incidence (30) of HCC and of death or transplantation in each score category, against model groups of the same size ranked on the predicted probability of HCC, and by the model’s predicted class (Gray’s test (31));
4. calibration and accuracy: the model’s probabilities rank patients but are not absolute risks (it was trained with class weights, eq. 3). We recalibrated them with a Fine–Gray model (32) on the two log-odds against event-free, cross-fitted, and gave each score the same treatment; we report the Brier score and the index of prediction accuracy (IPA) (33).

Because a first measurement may be ordered when HCC is already suspected, all analyses except calibration were repeated after a 6-month washout (patients with an event or censoring in the first 6 months excluded, time counted from the end of the washout). We report medians over the 20 repeats and, for the AUC( ) and concordance differences, the median of each per-repeat CI bound. Subgroups (sex, age, etiology, prevalent or incident cirrhosis, and the two of the sensitivity analyses below) are in Supplementary Table 3; an AUC( ) was not computed with fewer than 10 HCC cases. The design follows the recommendations for validating prediction models under competing risks (34), and the reporting follows TRIPOD+AI (35) (checklist in the supplementary material); Supplementary Table 14 lists the design safeguards against the biases of routine data.

### Deployment model and absolute-risk recalibration

The deployment model was trained on the whole development set (17,177 patients). Its recalibration to absolute risk was fitted on routine laboratory visits, the first per patient in each 6-month window from 2020 to 2024 (82,134 visits), stacked, with features rebuilt at each visit and out-of-fold predictions (a landmark supermodel for the recalibration only (36)). The predicted risk of HCC by month . is

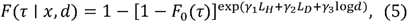

with *L_H_* and *L_D_* the log-odds of HCC and of death or transplantation against event-free, *d* the years since the cirrhosis record, γ, = (1.053,—0.283,0.106) and *F*_0_ (6,12,24,36) - 0.0099,0.0170,0.0296,0.0418 (medians over 20 repeats; Supplementary Table 6). Because both log-odds are sums over features (eq. 2), the deployment model’s absolute risk is itself additive on the subdistribution-hazard scale:

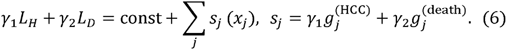

Each feature’s value multiplies the subdistribution hazard of HCC by exp(*s_j_*) (Supplementary Fig. 9). γ_2_ < 0: a death signal lowers the incidence of HCC, as a competing risk does.

The deployment model is stored as a JSON file with a numpy-only reference scorer, which also returns each feature’s contribution to a prediction. The file holds 92,988 bin scores (110 terms × 3 classes), 3 coefficients and 4 baseline values. A new health system would refit *F_0_*, then γ.

### Sensitivity analyses

Four further analyses addressed the main threats to these conclusions. First, to approximate validation in a new region, we used internal–external cross-validation by province (37): the reference model was refitted eight times, each on the development patients of seven of the eight Andalusian provinces (assigned from the patient’s primary-care health district), with the same pipeline, hyperparameters and class weights as the deployment model, and applied to the eighth. Each held-out province was evaluated in the development design, against the cross-validated predictions for the same patients, and at the score landmarks, where the province-level differences in concordance were pooled with a DerSimonian–Laird random-effects model (38). Second, because laboratory tests are ordered by clinicians, we compared the model with the scores separately in patients with all four key tests (AFP, platelets, albumin and bilirubin) measured in the history window and in those with at least one missing, and by calendar year of the landmark. Third, the visit-based recalibration has no sex term; we refitted it with a female indicator, cross-fitted by fold, and compared observed/expected ratios by sex. Fourth, because the recalibration was fitted at laboratory visits, we applied it at time zeros that need not be visits: each anniversary of the study start (2 January 2020 to 2024), to every patient eligible on that date, with features rebuilt from the previous 12 months and out-of-fold predictions, and compared observed/expected ratios by whether a laboratory visit fell in the preceding 6 months.

### OMOP standardisation

Source data were transformed into the Observational Medical Outcomes Partnership (OMOP) Common Data Model (CDM) (39,40). Because the entire machine-learning pipeline was built against the OMOP standard rather than against dataset-specific logic, other researchers can reproduce the same process on their own data once it is likewise standardised to OMOP CDM, improving the reproducibility of the overall methodology.

Unlike OMOP, which organises data by clinical domain (conditions, drugs, measurements, etc.), the BPS source tables were organised by their originating system or department. To bridge this difference, table column names were first harmonised: identifier columns were mapped to OMOP’s person_id, start and end date columns were renamed to generic names, and a type_concept_id was defined for each table. After this preprocessing, each OMOP clinical table could be built on top of this pattern, supplemented with any additional records specific to that table.

The Person and Death tables were directly derived from patients’ sociodemographic information. Location and Provider tables were also generated for completeness, although they were not used in the training or evaluation.

The Condition Occurrence and Procedure Occurrence tables were generated from condition-related and procedure-related tables, respectively. These tables contained exclusively ICD-9 and ICD-10 codes, which can be systematically mapped to standard codes in the OMOP Vocabularies ecosystem.

Drug occurrence was built from hospitals and outpatient pharmacy records. These identify each medication with a Spanish National Code, assigned by the Spanish Agency for Medicines and Health Products (AEMPS). Each code maps to an ATC code and a Virtual Therapeutic Moiety (VTM) code, both of which are represented within the OMOP Vocabularies repository, allowing these codes to be mapped directly.

Measurement records use the Andalusian CLC (Código de Laboratorio Clínico) system, a standardised nomenclature designed by the “Grupo de Nomenclatura y Codificación” (GNC) of the Andalusian Public Health Service to uniquely identify clinical laboratory tests and measurements. Most CLC codes map to LOINC codes, present in the OMOP Vocabularies, or to IUPAC nomenclature.

The local vocabularies employed by the source data were registered as custom codes in the Concept table, with their corresponding vocabularies added to the Vocabulary table. Whenever possible, a plausible code was manually assigned to any code that could not directly be mapped to a standard OMOP code, registering the mapping in both the Concept Relationship and Source To Concept Map OMOP tables. This ensured that a record for every mapping decision was kept in case a future revision is needed.

Finally, the Visit Occurrence and Observation Period tables were derived from all the other tables combined, since each record of the source tables reflects an interaction between the patient and the health system.

### Software stack

All analyses were conducted using Python 3.10.18 (41). The main machine learning and data processing libraries included scikit-learn 1.7.1 (42) for model training and evaluation, pandas 2.3.1 (43) and polars 1.31.0 (44) for data manipulation, scipy 1.15.2 (45) with numpy 2.2.6 (46) for numerical operations, interpret 0.6.12 for Explainable Boosting Machines (25), comprisk 0.8.0 for competing-risks analysis (47), tableone 0.9.5 (48) for descriptive tables, joblib 1.5.1 (49) for parallel computing, and matplotlib 3.10.3 (50) for visualisation. For reproducibility, we used pixi 0.70.0 (51) for dependency management and environment configuration, and DVC 3.61.0 (52) for pipeline management and version control (data and model artifacts).

All code and scripts used for data analysis are available at https://github.com/Computational-Medicine-Platform-FPS/bps-pred-hcc.

## Results

### Study population

Of 30,736 adults with cirrhosis, 17,177 had an eligible prediction date between 2020 and 2023 and formed the development set (Fig. 1 b). Within 12 months of the prediction date, 771 (4.5%) developed HCC, 3,314 (19.3%) died or underwent liver transplantation without HCC (3,061 deaths and 253 transplants) and 13,092 (76.2%) remained event-free (Table 1). Women accounted for 34.9% of the development set but 17.3% of HCC cases. Patients who died without HCC were older and had lower albumin concentrations, those who underwent transplantation were younger and had the most advanced liver disease, and those who developed HCC had lower platelet counts and the highest AFP concentrations. In the 12-month history window, 79% of patients had a platelet count and 41% an AFP measurement.

**Table 1.**
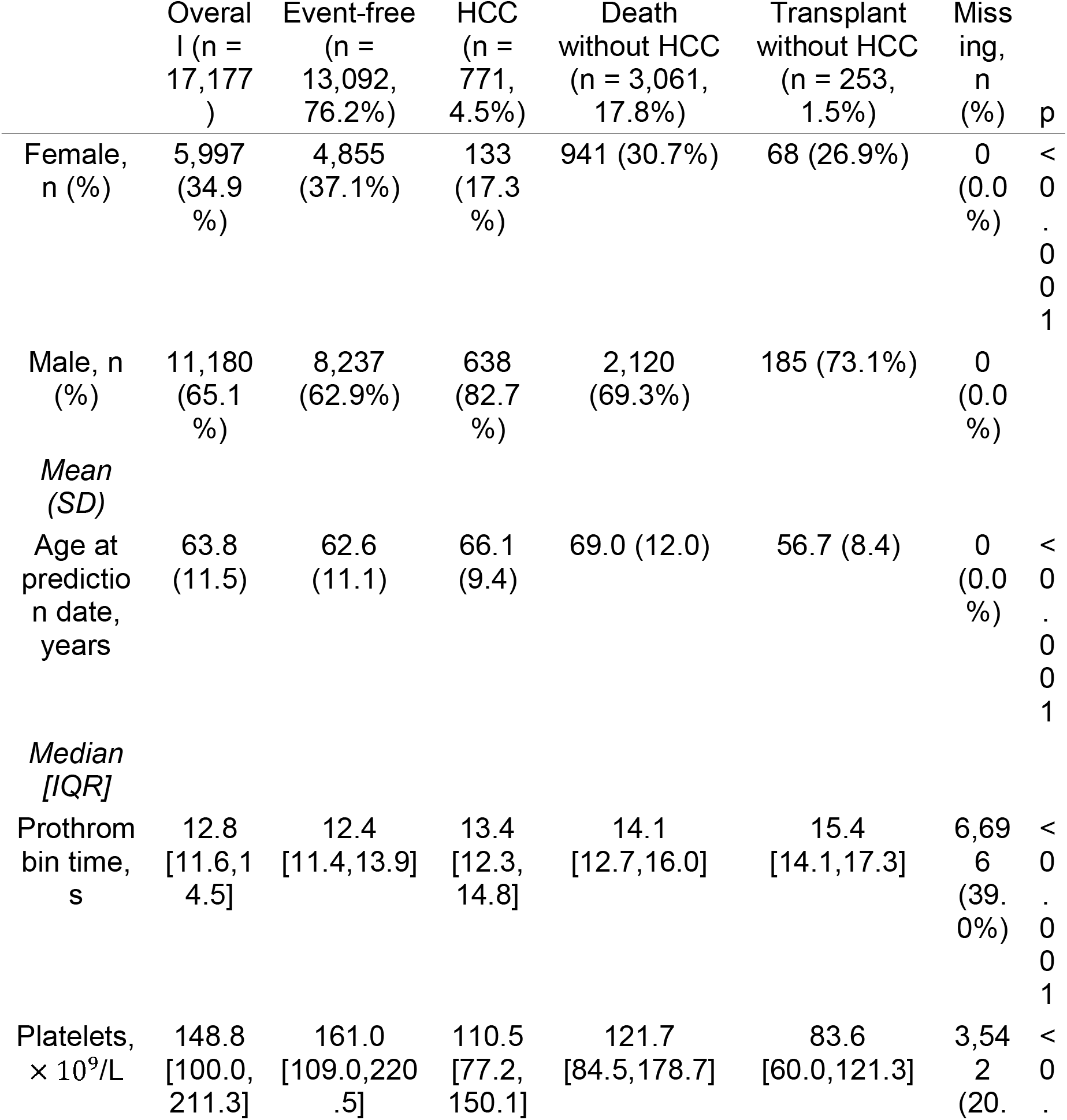
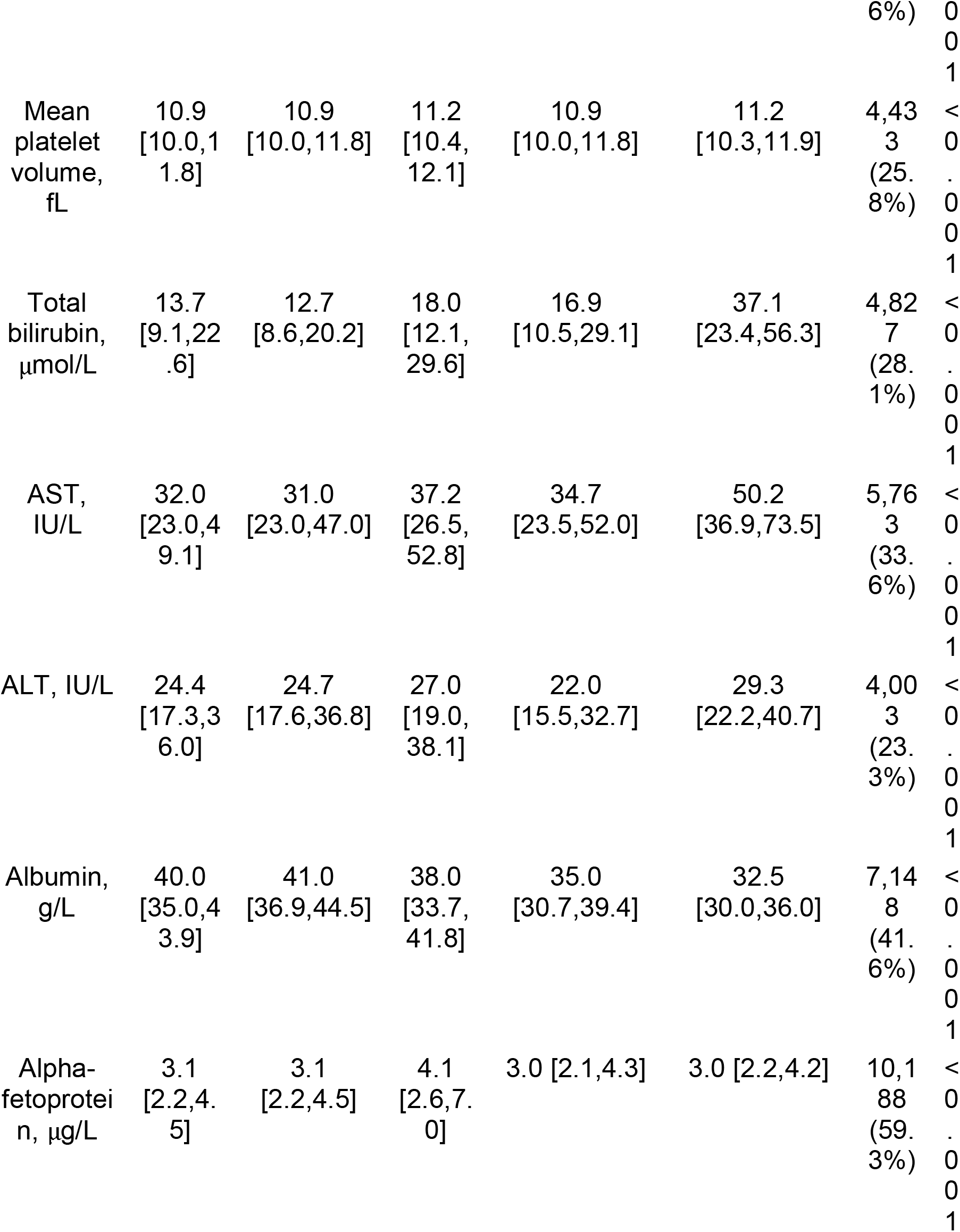
Characteristics of the development set by outcome within 12 months of the prediction date. Death and transplant form one class in the model and are shown separately here. Age is at the prediction date. Laboratory values are each patient’s mean over the 12-month history window, summarised as median [IQR]; missing means no measurement in that window. p values: χ^2^ test for sex, one-way ANOVA for age and Kruskal–Wallis test for laboratory values, across the four outcome groups. No cell holds fewer than 10 patients.

|  | Overall (n = 17,177) | Event-free (n = 13,092, 76.2%) | HCC (n = 771, 4.5%) | Death without HCC (n = 3,061, 17.8%) | Transplant without HCC (n = 253, 1.5%) | Missing, n (%) | p |
|---|---|---|---|---|---|---|---|
| Female, n (%) | 5,997 (34.9%) | 4,855 (37.1%) | 133 (17.3%) | 941 (30.7%) | 68 (26.9%) | 0 (0.0%) | <0.001 |
| Male, n (%) | 11,180 (65.1%) | 8,237 (62.9%) | 638 (82.7%) | 2,120 (69.3%) | 185 (73.1%) | 0 (0.0%) |  |
| *Mean (SD)* |  |  |  |  |  |  |  |
| Age at prediction date, years | 63.8 (11.5) | 62.6 (11.1) | 66.1 (9.4) | 69.0 (12.0) | 56.7 (8.4) | 0 (0.0%) | <0.001 |
| *Median [IQR]* |  |  |  |  |  |  |  |
| Prothrombin time, s | 12.8 [11.6,14.5] | 12.4 [11.4,13.9] | 13.4 [12.3, 14.8] | 14.1 [12.7,16.0] | 15.4 [14.1,17.3] | 6,696 (39.0%) | <0.001 |
| Platelets, × 10<sup>9</sup>/L | 148.8 [100.0,211.3] | 161.0 [109.0,220.5] | 110.5 [77.2, 150.1] | 121.7 [84.5,178.7] | 83.6 [60.0,121.3] | 3,542 (20.6%) | <0.001 |
| Mean platelet volume, fL | 10.9 [10.0,11.8] | 10.9 [10.0,11.8] | 11.2 [10.4, 12.1] | 10.9 [10.0,11.8] | 11.2 [10.3,11.9] | 4,433 (25.8%) | <0.001 |
| Total bilirubin, µmol/L | 13.7 [9.1,22.6] | 12.7 [8.6,20.2] | 18.0 [12.1, 29.6] | 16.9 [10.5,29.1] | 37.1 [23.4,56.3] | 4,827 (28.1%) | <0.001 |
| AST, IU/L | 32.0 [23.0,49.1] | 31.0 [23.0,47.0] | 37.2 [26.5, 52.8] | 34.7 [23.5,52.0] | 50.2 [36.9,73.5] | 5,763 (33.6%) | <0.001 |
| ALT, IU/L | 24.4 [17.3,36.0] | 24.7 [17.6,36.8] | 27.0 [19.0, 38.1] | 22.0 [15.5,32.7] | 29.3 [22.2,40.7] | 4,003 (23.3%) | <0.001 |
| Albumin, g/L | 40.0 [35.0,43.9] | 41.0 [36.9,44.5] | 38.0 [33.7, 41.8] | 35.0 [30.7,39.4] | 32.5 [30.0,36.0] | 7,148 (41.6%) | <0.001 |
| Alpha-fetoprotein, µg/L | 3.1 [2.2,4.5] | 3.1 [2.2,4.5] | 4.1 [2.6,7.0] | 3.0 [2.1,4.3] | 3.0 [2.2,4.2] | 10,188 (59.3%) | <0.001 |

### Cross-validated discrimination was not improved by interactions, a binary outcome or the full health record

In repeated cross-validation on outcome-anchored prediction dates, the reference model discriminated 12-month HCC with an AUROC of 0.734 ± 0.018 and an AUPRC of 0.148, approximately three times the outcome prevalence (0.045), and death or transplantation with an AUROC of 0.759 (Table 2). The training AUROC was moderately higher (0.794), indicating limited overfitting. At a threshold set for a 10% false-positive rate, the model identified approximately one third of the patients who developed HCC within a year (sensitivity 0.31; PPV 0.13; NPV 0.97).

**Table 2.** Development performance: the reference model and three variants on the same 100 cross-validation folds. Mean ± SD over the 100 test folds (20 repeats × 5 folds), with the same patients and folds for every model, on outcome-anchored prediction dates (development design). ΔAUROC: Nadeau–Bengio corrected paired t-test, Bonferroni-adjusted over the three comparisons. Probabilities: Bayesian correlated t-test with a region of practical equivalence of ± 0.01. HCC prevalence within 12 months: 0.045. Reference model at the threshold set for a 10% false-positive rate on the validation splits: sensitivity 0.310 ± 0.040, specificity 0.905 ± 0.009, PPV 0.134 ± 0.015, NPV 0.965 ± 0.002, positive likelihood ratio 3.30. Training-split AUROC was 0.794 ± 0.007, against 0.734 on validation and test. The per-class metrics of every model are in Supplementary Table 2.

| Model | HCC AUROC | HCC AUPRC | Death/transplant AUROC | ΔAUROC vs reference (adjusted p) | P(better / equivalent / worse) |
|---|---|---|---|---|---|
| **Reference**: three classes, 110 features, no interactions | 0.734 ± 0.018 | 0.148 ± 0.021 | 0.759 ± 0.008 | – | – |
| Binary (HCC against the rest) | 0.741 ± 0.017 | 0.146 ± 0.020 | – | +0.007 (0.42) | 0.25 / 0.75 / 0.00 |
| With pairwise interactions | 0.732 ± 0.017 | 0.144 ± 0.022 | 0.764 ± 0.008 | -0.001 (1.00) | 0.00 / 1.00 / 0.00 |
| Full health record (3,000+ features) | 0.710 ± 0.020 | 0.130 ± 0.018 | 0.773 ± 0.008 | -0.024 (0.0016) | 0.00 / 0.02 / 0.98 |

None of the three variants improved HCC discrimination. The binary model and the model with pairwise interactions were practically equivalent to the reference (posterior probability of equivalence 0.75 and 1.00), and the shape functions of the model with interactions were indistinguishable from those of the reference, as main effects are fitted before interactions. The full health record, comprising more than 3,000 features, reduced HCC AUROC by 0.024 while increasing the AUROC for death or transplantation from 0.759 to 0.773, consistent with comorbidity and frailty features that predict mortality diluting the HCC-specific signal.

### The model ranked HCC better than aMAP and THRI at routine encounters

To estimate performance at the point of care, the cross-validated models were applied without retraining at the first routine encounter at which each score could be computed. The aMAP landmark set comprised 14,928 patients, of whom 791 developed HCC during follow-up, and the THRI landmark set 18,848 patients, with 975 HCC cases. The model discriminated HCC better than both scores at all horizons from 6 to 36 months (Fig. 2 a, d; Table 3). At 12 months, AUC(12 mo) was 0.790 for the model and 0.688 for aMAP (ΔAUC(12 mo) +0.102, 95% CI +0.071 to +0.132), and 0.761 and 0.703 for THRI (+0.057, +0.032 to +0.081).

**Fig. 2.**
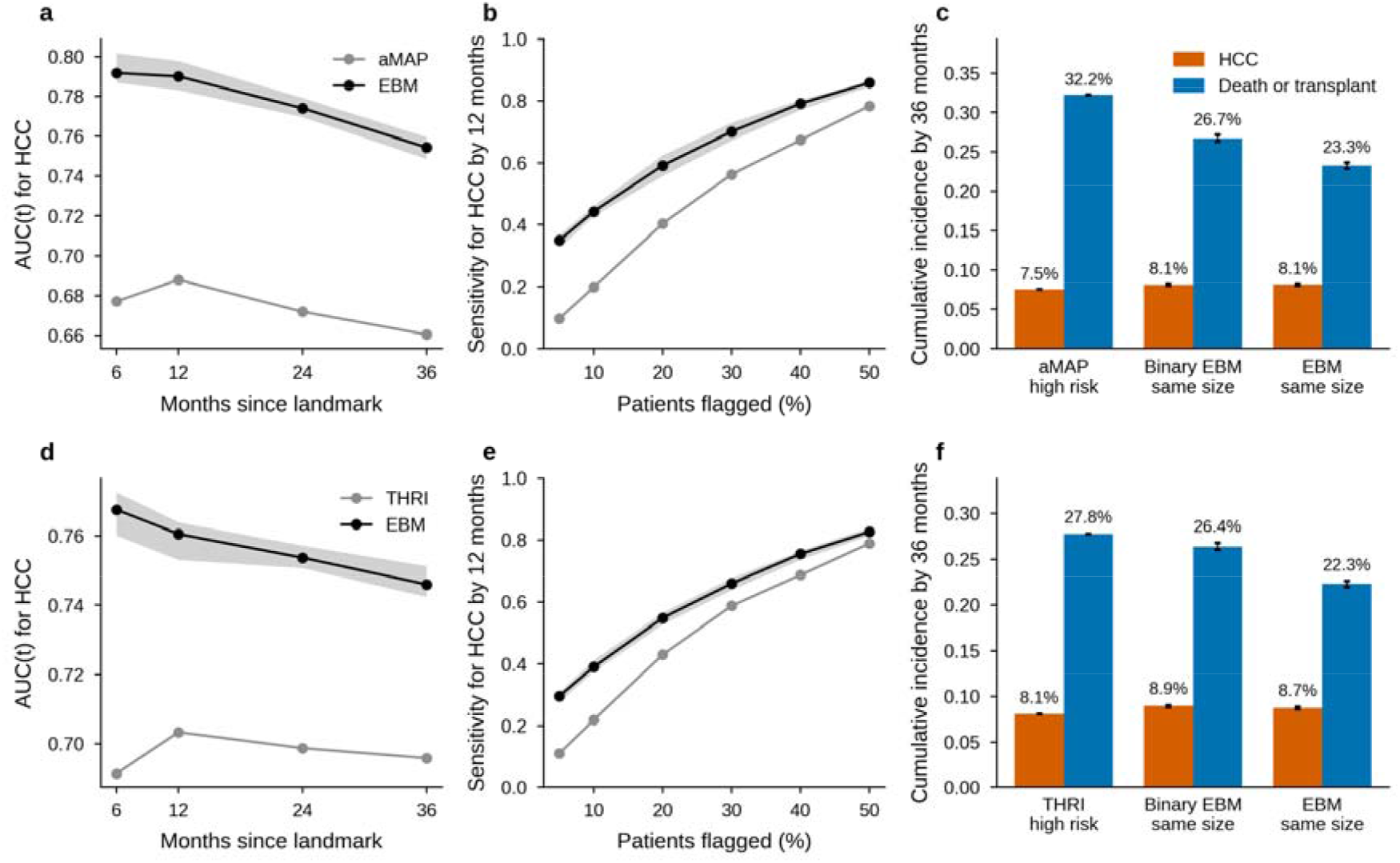
Discrimination of HCC at 6 to 36 months against aMAP and THRI, and composition of the high-risk groups. Top row: aMAP landmark set (14,928 patients, 791 HCC, 3,492 deaths or transplants during follow-up). Bottom row: THRI landmark set (18,848; 975; 4,547). Each patient is predicted by the cross-validation model that did not train on them (20 repeats). Lines and bars are medians over the repeats; bands and error bars span their range, i.e. the variability from model training. **a**, **d**, Time-dependent AUC for HCC, AUC( ) (cumulative/dynamic, inverse probability of censoring weights), with deaths and transplants by counted as negatives, for the model (black) and the score (grey); the paired differences and their 95% CIs are in Table 3. **b**, **e**, Sensitivity for HCC diagnosed within 12 months when each marker flags the same share of patients, estimated as qF̂_flagged_(12)/F̂_all_(12) from Aalen–Johansen estimates; at 10% flagged: model 44% vs aMAP 20%, model 39% vs THRI 22%. **c**, **f**, Aalen–Johansen cumulative incidence of HCC (vermilion) and of death or liver transplantation without HCC (blue) by 36 months in three groups of the same size: the score’s high-risk category (aMAP≥ 60: 59.3% of the landmark set; THRI > 240: 42.9%), the patients the binary EBM ranks highest, and the patients the three-class model ranks highest on the probability of HCC. All three hold about as much HCC. Deaths or transplants per case of HCC were 4.3 (score), 3.3 (binary EBM) and 2.9 (EBM) in the aMAP landmark set, and 3.4, 2.9 and 2.6 in the THRI landmark set.

**Table 3.** Comparison with aMAP and THRI at each score’s landmark. AUC(t): time-dependent AUC for HCC at t months (cumulative/dynamic, IPCW), not the time-fixed AUROC of Table 2. ΔC: difference in cause-specific concordance to 36 months. Medians over 20 repeats. CIs are the medians of the per-repeat bounds from 200 paired bootstrap resamples (AUC(t)) or influence functions (ΔC). High-risk group: the score’s high-risk category against the model’s group of the same size, with the 36-month cumulative incidence of HCC and of death or transplantation. IPA, index of prediction accuracy after the same cross-fitted Fine–Gray recalibration (not computed after the washout). O/E, observed/expected.

|  | aMAP | aMAP, 6-month washout | THRI | THRI, 6-month washout |
|---|---|---|---|---|
| Patients | 14,928 | 12,803 | 18,848 | 16,547 |
| HCC | 791 | 567 | 975 | 743 |
| Death/transplant | 3,492 | 2,456 | 4,547 | 3,441 |
| AUC(6 mo), score | 0.677 | 0.730 | 0.691 | 0.727 |
| AUC(6 mo), EBM | 0.792 | 0.783 | 0.768 | 0.749 |
| ΔAUC(6 mo) (95% CI) | +0.115 (+0.075, +0.155) | +0.053 (+0.005, +0.099) | +0.076 (+0.044, +0.110) | +0.022 (-0.016, +0.057) |
| AUC(12 mo), score | 0.688 | 0.721 | 0.703 | 0.724 |
| AUC(12 mo), EBM | 0.790 | 0.762 | 0.761 | 0.748 |
| ΔAUC(12 mo) (95% CI) | +0.102 (+0.071, +0.132) | +0.041 (+0.010, +0.077) | +0.057 (+0.032, +0.081) | +0.024 (-0.009, +0.053) |
| AUC(24 mo), score | 0.672 | 0.701 | 0.699 | 0.709 |
| AUC(24 mo), EBM | 0.774 | 0.752 | 0.754 | 0.746 |
| ΔAUC(24 mo) (95% CI) | +0.102 (+0.079, +0.128) | +0.052 (+0.021, +0.080) | +0.055 (+0.039, +0.074) | +0.037 (+0.014, +0.056) |
| AUC(36 mo), score | 0.660 | 0.676 | 0.696 | 0.699 |
| AUC(36 mo), EBM | 0.754 | 0.734 | 0.746 | 0.738 |
| ΔAUC(36 mo) (95% CI) | +0.094 (+0.071, +0.114) | +0.058 (+0.036, +0.081) | +0.050 (+0.036, +0.067) | +0.039 (+0.018, +0.058) |
| ΔC to 36 mo (95% CI) | +0.085 (+0.064, +0.106) | +0.055 (+0.032, +0.078) | +0.048 (+0.031, +0.065) | +0.040 (+0.022, +0.059) |
| High-risk group size (%) | 59.3% | 57.3% | 42.9% | 42.0% |
| High-risk 36 mo HCC / death, score | 7.5% / 32.2% | 7.1% / 29.6% | 8.1% / 27.8% | 7.7% / 26.3% |
| High-risk 36 mo HCC / death, EBM | 8.1% / 23.3% | 7.6% / 22.4% | 8.7% / 22.3% | 8.3% / 22.1% |
| IPA 12 mo, score (recalibrated) [range] | 0.8% [0.8%, 0.8%] | – | 1.0% [0.9%, 1.0%] | – |
| IPA 12 mo, EBM (recalibrated) [range] | 9.5% [8.0%, 10.0%] | – | 6.3% [5.2%, 7.0%] | – |
| Visit-based recalibration, 12 mo O/E; slope | 1.12; 1.02 | – | 1.05; 1.01 | – |

Because some early diagnoses may follow investigations prompted by suspicion of HCC, the analyses were repeated after a 6-month washout; the advantage was attenuated but persisted, with differences in concordance to 36 months of +0.055 against aMAP and +0.040 against THRI, both with confidence intervals excluding zero. When each marker flagged 10% of patients, the model identified approximately twice as many of the HCC cases diagnosed within 12 months (44% vs 20% for aMAP; 39% vs 22% for THRI; Fig. 2 b, e).

After identical recalibration, the index of prediction accuracy for 12-month HCC was 9.5% (aMAP landmark set) and 6.3% (THRI landmark set) for the model, compared with approximately 1% for either score (Supplementary Fig. 5), and the visit-based recalibration remained close to calibrated at both landmarks (observed/expected ratios 1.12 and 1.05). Results were consistent when hepatitis C was scored as cured in THRI, which reduced the score’s discrimination (Supplementary Fig. 2). The advantage was evident in men and largest in viral hepatitis, but was not significant in women (ΔC +0.029 against aMAP, 95% CI - 0.022 to +0.080), in whom the recalibration also overestimated risk, and was absent in MASLD (Supplementary Fig. 3; Supplementary Table 3).

### High-risk groups selected by the model contained fewer competing deaths

For surveillance, the composition of a high-risk group matters as well as the number of cancers it contains. In groups of the same size as each score’s high-risk category, the model’s group contained a similar proportion of HCC cases but fewer patients who died or underwent transplantation first: 2.9 competing events per HCC case, compared with 4.3 in the aMAP high-risk category, and 2.6 compared with 3.4 for THRI (Fig. 2 c, f; Supplementary Fig. 1). The binary model ranked 12-month HCC slightly better than the three-class model (three-class minus binary, AUC(12 mo) -0.019 and -0.021; C to 36 months -0.006 and -0.007), but its high-risk group contained more competing events (26.7% vs 23.3%) and its recalibrated risks were less accurate (index of prediction accuracy 7.8% vs 9.5%; Supplementary Fig. 4).

### Shape functions separated cancer signals from markers of liver failure

Because the model is a sum of shape functions, its behaviour can be examined directly. All values reported here are contrasts against remaining event-free, expressed as changes in log-odds and averaged over the 100 cross-validation models (Fig. 3; Supplementary Figs 6–9).

**Fig. 3.**
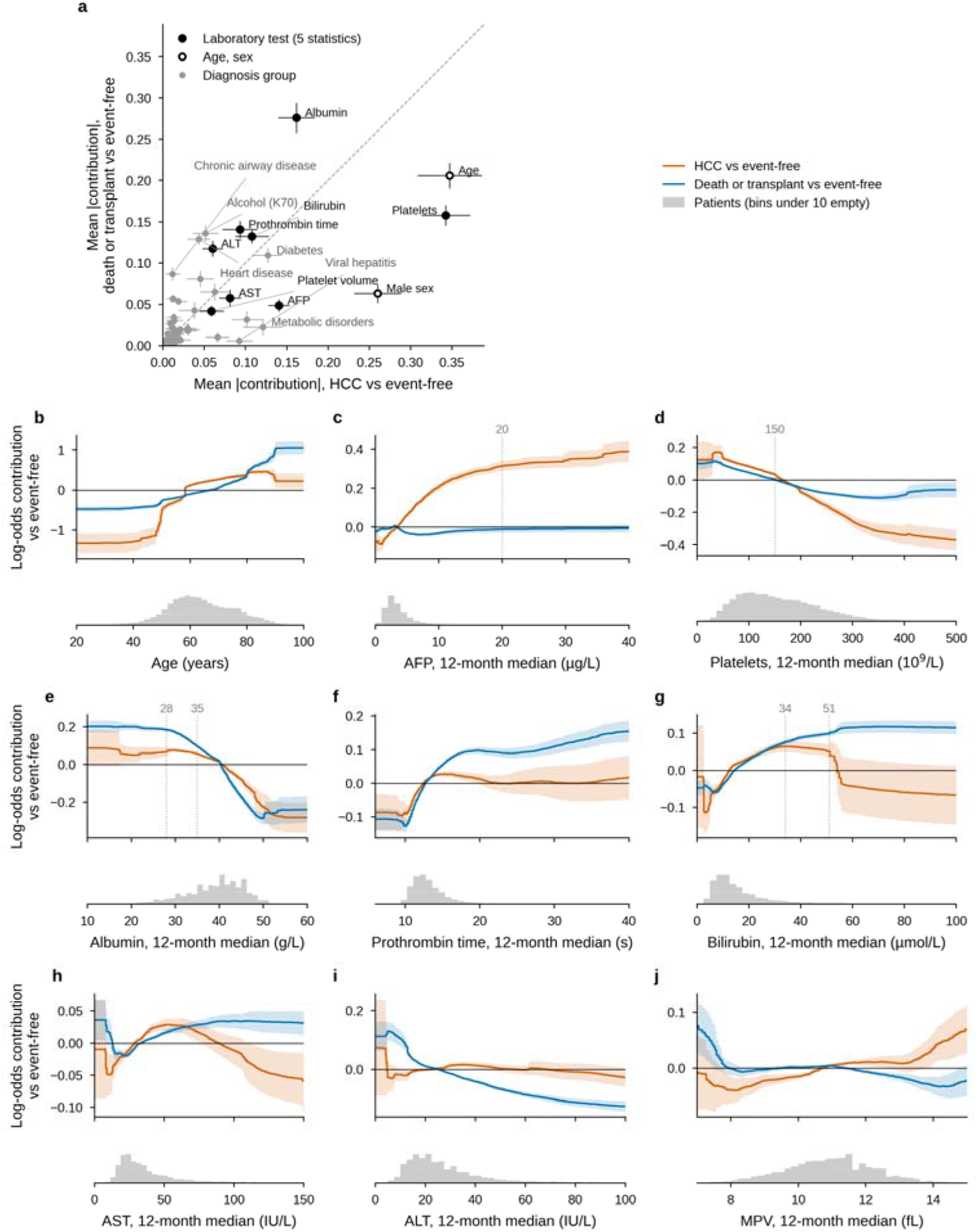
The model’s shape functions separate the cancer signal from liver failure. **a**, Signal map. For each feature group: the mean absolute contribution, over each model’s training patients, to the log-odds of HCC against event-free (x) and of death or liver transplantation against event-free (y). A laboratory test’s five statistics (mean, median, SD, maximum, minimum of the 12-month window) are summed per patient first. Error bars are the SD over the 100 cross-validation models; the dashed line marks equal importance for both contrasts. AFP, platelets, male sex and viral hepatitis lie below the diagonal (HCC); albumin, prothrombin time, ALT, alcohol-related liver disease and cardiorespiratory disease lie above it (death or transplant); age and bilirubin contribute to both. **b–j**, Shape functions of age (b) and of the 12-month median of each of the eight laboratory tests: AFP (c), platelets (d), albumin (e), prothrombin time (f), bilirubin (g), AST (h), ALT (i) and mean platelet volume (MPV, j). Each panel shows the change in the log-odds of HCC (vermilion) and of death or transplantation (blue) against staying event-free, as fold mean ±SD; every curve is centred to mean zero over the training patients. Reference lines mark AFP 20 μg/L, platelets 150 × 10^9^/L, and the Child–Pugh bands of albumin (28 and 35 g/L) and bilirubin (34 and 51 μmol/L, i.e. 2 and 3 mg/dL). Grey strips: patients per bin, bins under 10 left empty. A three-class model’s curves are identified only as contrasts against a reference class (Methods), hence two curves per panel. Binary model: Supplementary Fig. 7; binary features: Supplementary Fig. 8.

At the level of feature groups, predictors fell on either side of the line of equal contribution (Fig. 3 a). AFP, platelet count, male sex and viral hepatitis contributed more to the log-odds of HCC, whereas albumin, prothrombin time, ALT, alcohol-related liver disease (K70), heart disease and chronic airway disease contributed more to the log-odds of death or transplantation; age, bilirubin and diabetes contributed to both.

AFP was the most HCC-specific variable: it increased the log-odds of HCC steeply up to 20 µg/L (+0.31), without affecting the death contrast, and its within-year variability contributed an additional HCC-specific signal (Supplementary Fig. 6). Low platelet counts increased both contrasts, the HCC contrast more, with the effect changing sign near 165 × 10^9^ /L. The HCC contrast increased sharply between 48 and 50 years of age and plateaued between 82 and 88 years before declining, whereas the death contrast increased continuously and exceeded the HCC contrast from about 80 years.

Markers of hepatic reserve were associated predominantly with death. Albumin below 35 g/L increased the death contrast approximately three times more than the HCC contrast, and prolonged prothrombin time increased mainly the death contrast. Bilirubin increased the death contrast monotonically, whereas the HCC contrast decreased in a step between 51 and 55 µmol/L, at the boundary of the highest Child–Pugh bilirubin category (Fig. 3 g). This step was sharpest in the median term, the mean term declining gradually (Supplementary Fig. 6), and was learned from the data; it may reflect clinical practice at this threshold as well as biology (see Discussion). Very low ALT increased the death contrast, and AST above approximately 100 IU/L decreased the HCC contrast (Fig. 3 h, i). Male sex and viral hepatitis increased only the HCC contrast (+0.57 and +0.34), whereas K70, kidney failure and organic mental disorders increased the death contrast (Supplementary Fig. 8).

### The binary model concealed the competing-death signal

The shape functions of the binary model closely followed the HCC contrast of the three-class model, with the death signal incorporated at approximately one fifth of its weight. In the binary model, *h* = *g_HCC_* — *q g_death_* , where *q* = 0.20 is the proportion of deaths among patients without HCC (eq. 4); this approximation reproduced the binary curves (for albumin, a root-mean-square difference, weighted by the number of patients, of 0.004, compared with 0.030 for the HCC contrast alone; Supplementary Fig. 7). Consequently, a low value on a binary curve does not distinguish a low cancer risk due to preserved liver function from one due to impending death, a distinction that the three-class model makes explicit by displaying the death contrast alongside (Fig. 3).

### The deployment model gave calibrated absolute risks explained feature by feature

The deployment model returns, for each patient, every feature’s contribution to both log-odds and a recalibrated absolute risk. Two hypothetical patients with the same aMAP score (68.2, high risk) illustrate the output (Fig. 4 a–c). In a man with a cancer profile (AFP 15 µg/L, platelets 90 . 10^9^/L, preserved liver function), AFP and platelets dominated the HCC contrast, and the 12-month risk of HCC was 13.3%. In a man with a liver-failure profile (albumin 28 g/L, bilirubin 70 µmol/L), albumin and bilirubin dominated the death contrast, and the risk was 2.9%, 4.6 times lower.

**Fig. 4.**
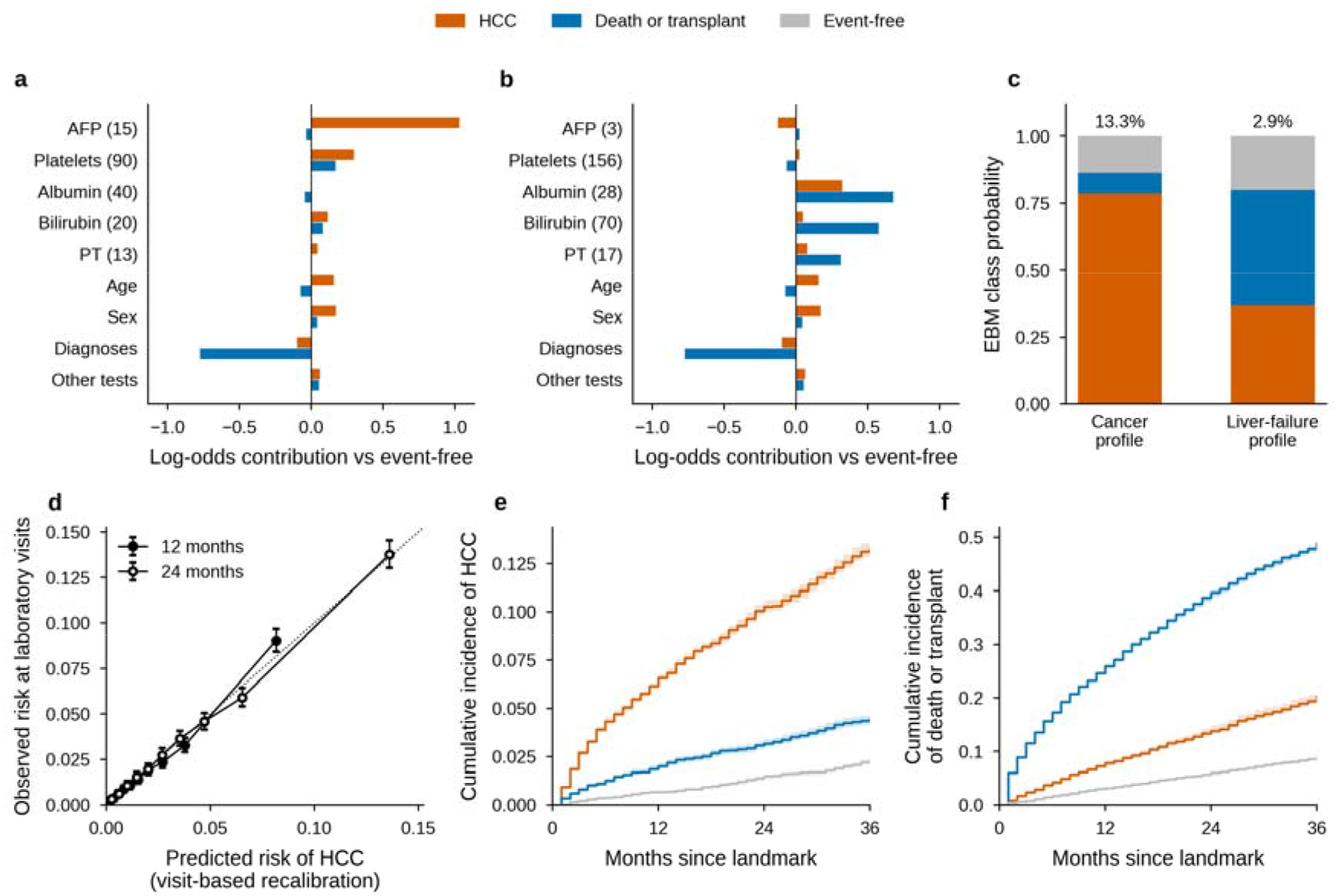
From shape functions to an absolute risk and a three-way triage. **a**, **b**, Two hypothetical men aged 65, both with a K74 cirrhosis code, three years after the cirrhosis record, with the same aMAP score of 68.2 (high risk): a cancer profile (a; AFP 15 g/L, platelets 90 /L, albumin 40 g/L, bilirubin 20 mol/L, PT 13 s) and a liver-failure profile (b; AFP 3 g/L, platelets 156 /L, albumin 28 g/L, bilirubin 70 mol/L, PT 17 s). Each test’s mean, median, minimum and maximum take the value shown; its SD, and any test not shown, take the training median. Bars are each feature group’s contribution to the log-odds of HCC (vermilion) and of death or transplantation (blue) against event-free, from the deployment model file and its numpy scorer; with the intercept they sum exactly to the model’s log-odds. **c**, The two patients’ class probabilities, coloured as in the legend above the figure; the number above each bar is the recalibrated 12-month risk of HCC. Cancer profile: P(HCC) 0.79, P(death or transplant) 0.08, 12-month risk 13.3%, THRI 286. Liver-failure profile: 0.37, 0.43, 2.9% (4.6 times lower), THRI 236. With no diagnosis groups other than K74 recorded, both patients’ comorbidity lowers their death log-odds (-0.77). **d**, Calibration of the visit-based recalibration at routine laboratory visits, at 12 months (filled) and 24 months (open): deciles of predicted risk against the Aalen–Johansen observed risk, medians over 20 repeats with 95% CIs, the recalibration refitted without each fold’s patients. Observed/expected 1.00 at 12 months and 1.01 at 24 months; calibration slope 0.99; IPA 6.8% at 12 months. **e**, **f**, Aalen–Johansen cumulative incidence of HCC (e) and of death or liver transplantation (f) by the model’s predicted class at the aMAP landmark: predicted HCC (vermilion, n = 3,310), predicted death or transplant (blue, n = 4,627), predicted event-free (grey, n = 6,996); medians over 20 repeats. By 36 months, 13.2% of the predicted-HCC class developed HCC (against 4.4% and 2.3% in the other classes), and 48.8% of the predicted-death class died or were transplanted (against 20.3% and 8.8%). Gray’s test p < 10^−70^ for each cause, in all 20 repeats.

Because the model was trained with class weights, its raw probabilities ranked patients correctly but overestimated risk (Brier score 0.111, compared with 0.043 for a model assigning every patient the prevalence). After visit-based recalibration, predicted and observed 12-month risks agreed at routine laboratory visits (observed/expected ratio 1.00; calibration slope 0.99; Fig. 4 d).

Because the recalibration is linear in the two log-odds (eq. 6), the shape functions translate directly into subdistribution hazard ratios: an AFP of 20 µg/L multiplied the subdistribution hazard of HCC by 1.37 and an age between 30 and 48 years by 0.27 to 0.32, whereas contributions to the death contrast, entering with γ_2_ < 0, reduced it (Supplementary Fig. 9).

At the population level, the predicted class separated the three outcomes at the aMAP landmark: by 36 months, 13.2% of patients predicted to develop HCC had done so, and 48.8% of those predicted to die or undergo transplantation had died or been transplanted (Fig. 4 e, f).

### Sensitivity analyses

Leaving each province out of training in turn did not lower discrimination. The held-out provinces’ AUROC for HCC was 0.738 (0.712 to 0.791 across provinces), against 0.733 for the cross-validated models in the same patients, and the advantage over the scores held, with pooled concordance differences of +0.091 (95% CI +0.068 to +0.114; I^2^ 23%) against aMAP and +0.057 (+0.040 to +0.073; I^2^ 0%) against THRI (Supplementary Table 10). The advantage was confined to patients with all four key laboratory tests measured in the history window (ΔC +0.090 against aMAP and +0.087 against THRI); with any missing, it was not significant (+0.029, 95% CI -0.015 to +0.074, and +0.013, -0.011 to +0.036). It was similar across landmark years (Supplementary Fig. 3; Supplementary Table 3).

At laboratory visits, the recalibration overestimated short-term risk in women (observed/expected 0.89 at 12 months, against 1.03 in men), converging by 36 months; a female term barely changed this (0.91), with a subdistribution hazard ratio of 0.98 (range over the refits 0.81 to 1.11; Supplementary Table 11). At calendar time zeros, the model still ranked patients (AUC(12 mo) 0.75, against 0.78 at visits; calibration slope 0.96), but the visit-based recalibration overestimated risk (observed/expected 0.79 at 12 months and 0.94 at 36 months), most in patients without a laboratory visit in the previous 6 months (0.68, against 0.85 with one; Supplementary Table 12).

## Discussion

### Principal findings

An interpretable three-class model, in which the effect of each variable is displayed as a curve, discriminated short-term HCC better than aMAP and THRI at routine encounters, at every horizon up to three years and after excluding HCC diagnosed within six months. It used only data already held in the health record (age, sex, coded diagnoses and eight routine laboratory tests). At the same group size, its high-risk group held as much HCC as the scores’ high-risk categories with 20–28% fewer patients who died or underwent transplantation first, and after recalibration at routine visits its absolute risks agreed with observed incidence. The advantage held when each province was left out of training. Neither interaction terms, nor the full health record, nor a binary formulation improved discrimination, and the binary formulation concealed the competing-death signal within its single curve.

### Clinical interpretation of the shape functions

Interpretable modelling allowed us to examine clinical hepatology heuristics directly in routine data. Instead of producing only opaque risk probabilities, the model showed how each biomarker shifted prediction toward HCC, death or transplantation without HCC, or event-free survival. This made it possible to separate signals of oncogenesis from signals of hepatic deterioration and competing mortality.

The shape functions followed clinically plausible patterns. We describe each as two curves: its contribution to the log-odds of HCC, and to the log-odds of death or transplantation, each relative to remaining event-free (the HCC and death contrasts). Low albumin, prolonged prothrombin time, and higher bilirubin increased the model contribution toward death without HCC, consistent with impaired liver reserve and with how albumin and bilirubin anchor the ALBI liver function grade (53). The bilirubin function also recovered a clinical threshold without being given it: the HCC contrast dropped in a step at 51 µmol/L, the boundary of the highest Child–Pugh bilirubin band, while the death contrast kept rising. This step may reflect disease biology, but also care: Child–Pugh C prompts transplant evaluation and, outside the transplant list, surveillance is no longer recommended (4), and in routine records the ordering and timing of tests carry information beyond their values (54). We therefore read the recovered thresholds as the joint footprint of disease and clinical practice, which is what a model deployed in the same health system will encounter.

AFP showed the most HCC-specific response among all evaluated biomarkers. The learned function preserved a continuous risk gradient rather than imposing a binary threshold: the HCC contribution rose steeply from 2 to 20 µg/L (+0.31 at 20 µg/L) and more slowly above it, with no death signal. This gradient spans the conventional threshold of 20 ng/mL (20 µg/L) used in HCC surveillance and recall strategies (55,56), and is compatible with recent proposals of a lower threshold (57). Longitudinal surveillance data also support this graded interpretation, showing that AFP can discriminate HCC before diagnosis, with reported AUROC values of 0.792 at 12 months, 0.823 at 6 months, and 0.867 at diagnosis (58). Its within-year variability added an HCC-only signal, so a rising or fluctuating AFP below conventional thresholds carried information. Overall, the model recovered AFP risk regions directly from real-world data while retaining clinically relevant subthreshold information.

Platelet count added a clinically coherent marker of portal hypertension and cirrhosis severity. The HCC contribution rose as platelets fell below about 165 × 10^9^/L, consistent with thrombocytopenia as a marker of the cirrhotic substrate in which HCC risk emerges, and the death contribution moved the same way but less. This pattern aligns with the clinical use of 150 × 10^9^/L in portal-hypertension risk stratification, including Baveno VII criteria (59,60), and with the inclusion of platelets in aMAP for HCC risk prediction (7). Unlike AFP, platelet count did not behave as a cancer-specific biomarker. Instead, it contextualised cirrhosis severity and competing risk.

Although some recent models use the AST/ALT ratio as a single feature (61), entering both separately let the model give them different roles. AST above about 100 IU/L lowered the HCC contrast, in line with the AST/ALT ratio tracking fibrosis and impaired liver function (62) and short-term mortality in viral cirrhosis (63). Very low ALT raised the death contrast, compatible with low aminotransferases as a marker of frailty. Unlike a population cohort in which elevated ALT predicted HCC (64) and Child A HCV cirrhosis, in which persistently elevated ALT did (65), ALT showed no clear HCC signal once the other markers were in the model.

Finally, age added a strong demographic gradient to the model interpretation. Younger patients were shifted toward event-free survival and away from HCC and death without HCC, consistent with age being an independent risk factor for HCC in cirrhosis, overall and in HCV-related cirrhosis (66), while with increasing age the contribution toward death or transplantation rose, in line with age predicting decompensation in compensated cirrhosis (67). The HCC contribution rose steeply between 48 and 50 and plateaued between 82 and 88, whereas the death contribution kept rising and exceeded it from about 80, which reflects competing mortality rather than a decline in intrinsic HCC risk. The three-class model thus separated age-related oncological risk from age-related mortality, a distinction that a binary formulation partly loses, since competing outcomes are naturally described as transitions between the clinical states of cirrhosis (68).

Liver-disease and comorbidity code profiles, encoded as binary features, behaved as clinical knowledge predicts. Viral hepatitis shifted prediction toward HCC with no death signal, consistent with viral cirrhosis being a principal HCC surveillance risk state (56). K70, alcohol-related liver disease, shifted prediction toward death rather than HCC: alcohol-associated cirrhosis carries clinically relevant HCC incidence (69) and alcohol-related HCC is often diagnosed late, with a worse prognosis (70); in our 12-month window the competing outcome dominated. Kidney failure, organic mental disorders, heart disease and chronic airway disease were associated with death. The model did not treat all codes as equivalent: it assigned cancer signal to viral etiology and male sex, and competing-death signal to alcohol and comorbidity.

### Value of the three-class formulation

Discrimination alone does not justify the three-class formulation, as the binary model ranked HCC almost as well. Its value lies in how it handles patients who die before developing HCC. Both scores lean on platelets and, in aMAP’s case, the albumin-bilirubin grade, which predict decompensation as much as cancer, so their high-risk categories fill with patients who die without HCC (aMAP: four deaths for every HCC). At the same size, the model’s high-risk group held as much HCC with 20–28% fewer competing deaths, the separation a surveillance programme needs, since a patient likely to die within months gains least from intensified screening. The binary model performed in between: its high-risk group contained more competing deaths (26.7% vs 23.3%), its recalibrated risks were less accurate (IPA 7.8% vs 9.5%), and its curves display only the cancer signal. The three-class model provides a similar ranking and displays both signals.

### Limits of the binary formulation

A binary model predicts HCC against all other patients, so patients who died without HCC are grouped with those who remained event-free. Its shape functions are therefore a blend: approximately the HCC curve of the three-class model minus one fifth of its death curve, one fifth being the average proportion of deaths among patients without HCC (eq. 4). Two consequences follow. First, the binary curves resemble the three-class HCC curves and discrimination is similar, so a binary model is not wrong about cancer. Second, the death signal is hidden within the same curve: where it is strong (low albumin, prolonged prothrombin time, old age), the binary curve bends downwards, so that a low binary risk may reflect a healthy liver or one about to fail, and the curve cannot show which (Supplementary Fig. 7). Interpretable models have exposed such mixtures before. In a widely cited example, a model of pneumonia mortality learned that asthma lowered the risk of death, because patients with asthma were admitted directly to intensive care and treated more aggressively (71); inspecting the model revealed the artefact. Here, modelling competing events as a separate outcome removes the mixture rather than only revealing it.

### Comparison with established scores and machine-learning models

Few HCC models, machine learning or regression, have been developed or compared under competing risks (1). In HCV cirrhosis, a random survival forest built under competing risks discriminated better than a Fine–Gray model (72). Nahon et al., in 4,867 patients from three prospective French cohorts under semiannual ultrasound surveillance, analysed under competing risks, found no advantage of a decision tree, a random survival forest or a deep neural network over aMAP and FASTRAK in C-index, Brier score or decision curves (2). Our setting differs in three ways that may explain the contrast. Their patients had compensated, biopsy-proven cirrhosis, three quarters of it after HCV cure and the rest controlled HBV or non-viral, under protocolised surveillance; ours are an unselected population of all etiologies and stages, with routine, irregular testing, where a model can use the full laboratory trajectory: the advantage was confined to patients whose key tests had all been measured in the preceding year (Supplementary Table 3). Their models used about fifteen demographic and laboratory variables measured once, at inclusion or at viral cure; ours used five summary statistics of each test over a year, and coded comorbidity. And our largest gain was in viral hepatitis, which they excluded unless controlled. They also censored liver transplantation, whereas we count it with death as the competing outcome. Part of our advantage lies in the first months after the landmark, when some diagnoses may follow tests ordered on suspicion; the 6-month washout is the conservative estimate, and it still favoured the model against both scores. Both scores were built for longer horizons (aMAP 5 years, THRI 10) than our three years; THRI’s etiology points depend on whether hepatitis C has been cured, which is not recorded, but scoring hepatitis C as active or as cured led to the same conclusion. Supplementary Table 13 compares recent models (10,73–76).

aMAP and THRI performed worse here than in most curated cohorts. aMAP’s pooled C-statistic is 0.81 in non-viral liver disease (1); after HCV cure, its C-index was 0.77 in a Scottish cohort and lower in a second UK cohort (77); and THRI’s external C was 0.77 (8). In Nahon et al.’s validation set, however, aMAP’s time-dependent AUC was 0.73 at 1 year and 0.70 at 3 years (2), close to ours. Our AUC( ) values for the scores, 0.66–0.70, come from an unselected routine-care population, with each score evaluated at the first encounter where it could be computed, over 6 to 36 months, and outcomes from coded records rather than protocolised surveillance. The comparison is paired on the same patients and the same follow-up, so these conditions apply to both markers.

A relevant recent machine-learning comparator using routine clinical data is PRE-Screen-HCC (11). However, the clinical target differs from ours. PRE-Screen-HCC was designed as a broad population-level prescreening tool, developed in UK Biobank (78) and externally tested in All of Us (79), with models evaluated in the full population and in a broader patients-at-risk subgroup that was not restricted to cirrhosis. It used random forest models explained post hoc with SHAP values, which approximate the contribution of each variable to a model that cannot itself be inspected, for a binary HCC versus no-HCC task; such explanations do not show whether a biomarker signals cancer risk or competing mortality. Our model instead targets a narrower surveillance problem: short-term HCC prediction among patients with established cirrhosis, while explicitly separating death or transplantation without HCC from event-free survival. This makes our three-class model less suited to broad population prescreening than PRE-Screen-HCC, but more informative for understanding short-term HCC risk, liver failure risk, and competing mortality in established cirrhosis.

### Implementation considerations

The model’s inputs are extracted automatically from the electronic health record: age, sex, coded chronic diagnoses and the results of eight laboratory tests over the previous 12 months. A test not performed in that window takes the training median, so it contributes as a typical value; age is required, and the reference scorer rejects a record without it. The model adds most where AFP, platelets, albumin and bilirubin have all been measured in the previous year (63% of the aMAP and 42% of the THRI landmark set; Supplementary Table 3), and its absolute risks are calibrated for use at a laboratory visit; at other dates they overestimate (Supplementary Table 12). No manual data entry or tuning is needed. The output, the risk of HCC by 6 to 36 months together with each feature’s contribution, is meant for the clinicians who decide on surveillance; a strong competing-death signal is a prompt to review liver function and goals of care, not an instruction.

### Limitations

This study has limitations. First, all evaluations were internal, in one regional health system and period. The landmark comparison applied the cross-validated models at routine encounters without retraining, and leaving each province out of training did not lower discrimination (Supplementary Table 10), but the provinces share one health service, laboratory network and coding practice. External validation, with local recalibration of at least the baseline risk, is needed before use elsewhere; the OMOP-standardised pipeline is meant to make it possible. Second, routine data limit what is measured: HCC was ascertained from diagnosis codes without review of imaging or histology, and sustained virological response after hepatitis C is not recorded, so THRI was computed under both assumptions. Testing is clinician-driven, and the advantage over the scores was confined to patients with the key tests measured (Supplementary Table 3): where testing is sparse, the model adds little, and a missing test enters as a median value. Third, the model showed no advantage in women, who were 34.9% of the development set but 17.3% of HCC cases, nor in MASLD, with few cases. In women the recalibration overestimated risk at short horizons, and a proportional sex term did not correct it (Supplementary Table 11). Fourth, the recalibration was fitted at laboratory visits and overestimated risk at time zeros that were not visits (observed/expected 0.79 at 12 months, 0.68 without a visit in the previous 6 months; Supplementary Table 12), so its absolute risks apply at a laboratory visit.

## Conclusion

An interpretable three-class model built on the routine records of a whole region ranked short-term HCC risk in cirrhosis better than aMAP and THRI under competing risks, and neither interaction terms, a binary outcome nor the full health record improved its discrimination. Modelling death or transplantation without HCC as a separate outcome kept patients who die first out of the high-risk group and gave shape functions that separate cancer from liver failure, a distinction that a binary model folds into one curve. Because the pipeline is standardised to the OMOP Common Data Model, the model can be recalibrated and validated in other health systems, the next step before it informs surveillance decisions.

## Data Availability

The datasets analyzed during the current study are available in the Population Health Database repository (Munoyerro-Muniz et al., 2020), subject to controlled access according to the regulation for the use of medical data for research in the Andalusian Health System. Patient-level data cannot leave the PAGEM secure environment. All code is available at http://github.com/Computational-Medicine-Platform-FPS/bps-pred-hcc.

## Contributors

**IG**: Software, Methodology, Writing - original draft, Writing - review & editing. **AR**: Conceptualization, Validation, Writing - review & editing. **LC-B**: Methodology, Validation, Writing - review & editing. **LC-B**: Methodology, Validation, Writing - review & editing. **JD**: Conceptualization, Resources, Funding acquisition, Validation, Writing - review & editing. **MR-G**: Conceptualization, Resources, Funding acquisition, Validation, Writing - review & editing. **CL**: Conceptualization, Formal Analysis, Software, Methodology, Supervision, Writing - original draft, Writing - review & editing. IG and CL accessed and verified the underlying data. All authors have read and agreed to the published version of the manuscript and had final responsibility for the decision to submit.

## Ethical considerations

This study used pseudonymised, retrospectively extracted data obtained through the regional data-governance framework established for secondary use of health information in Andalusia (14). The study protocol (“Desarrollo de un predictor temprano del Carcinoma Hepatocelular: Hacia una Detección Temprana en la Población de Andalucía”, with internal code SICEIA-2024-002905, protocol version v.5.06 from 25 October 2024) was reviewed and approved by the Ethics Committee for the Coordination of Biomedical Research in Andalusia (Acta 10/24), which issued a favourable opinion on 26 November 2024. Given the retrospective design and the pseudonymisation of the data at source, the Committee granted a waiver of informed consent for the Patient Information Sheet/Informed Consent. The study was conducted in accordance with the Declaration of Helsinki, the requirements of Spanish Law 14/2007 of 3 July on Biomedical Research, and Organic Law 3/2018 of 5 December on the Protection of Personal Data. Records were pseudonymised at source by the data-governance technical staff of the Andalusian Health Service before extraction, so researchers had no access to identifying information. The protocol was not entered in a public registry.

### Patient and public involvement

Patients and members of the public were not involved in the design, conduct, reporting or dissemination of this study, which used only retrospective, pseudonymised records, with no interaction between researchers and patients.

### Consent for publication

Not applicable.

### Data and code availability

The datasets analyzed during the current study are available in the Population Health Database repository (Muñoyerro-Muñiz et al., 2020), subject to controlled access according to the regulation for the use of medical data for research in the Andalusian Health System. Patient-level data cannot leave the PAGEM secure environment. All code is available at http://github.com/Computational-Medicine-Platform-FPS/bps-pred-hcc.

### Use of AI-assisted tools

During the preparation of this work, the authors used Claude Code (Anthropic) and Gemini (Google) to assist with code, drafting and English editing. The authors reviewed and verified all outputs and take full responsibility for the content.

## Acknowledgments

This work was supported by grants PID2023_152380OB and PRTR-C17.I1 from the Spanish Ministry of Science and Innovation; grant PMP24/00005 from the Instituto de Salud Carlos III (ISCIII); grant IE19_259 FPS from Consejería de Salud y Consumo, Junta de Andalucía; and Horizon Europe (HORIZON), Marie Skłodowska-Curie Actions Doctoral Networks (101226456-MLCARE). The funders had no role in study design, data collection, data analysis, data interpretation or writing of the report.

