## Supplementary for "Interpretable competing-risk prediction of hepatocellular carcinoma in cirrhosis from region-wide routine health data"

#### Supplementary Methods: mathematical details

1. **Features.** For laboratory test $\ell$ with results $v_1, \dots, v_m$ in $[t_0 - 365\,\mathrm{d}, t_0)$: $x_{\ell,s} = s(v_1, \dots, v_m)$, $s \in \{\mathrm{mean}, \mathrm{median}, \mathrm{SD}, \mathrm{max}, \mathrm{min}\}$. A missing value takes the training

median, after robust scaling (which binning makes immaterial).

2. **Model.** $\eta_k(x) = \beta_k + \sum_j f_{jk}(x_j)$, $P(Y = k \mid x) = e^{\eta_k} / \sum_l e^{\eta_l}$, $k \in \{0, 1, 2\}$ (main eq. 1). Each $f_{jk}$ is constant within the bins of feature $j$ (bin 0 for missing). It is fitted by cyclic gradient boosting with bagging (interpret 0.6.12; every hyperparameter and its value in Supplementary Table 8) and centred per class with the training weights.

3. **Identifiability.** For any functions $c_j$, replacing $f_{jk}$ by $f_{jk} + c_j(x_j)$ for all $k$ leaves $P$ unchanged. Contrasts $g_j^{(k)} = f_{jk} - f_{j0}$ are invariant, and $\log[P_k/P_0] = (\beta_k - \beta_0) + \sum_j g_j^{(k)}(x_j)$ (main eq. 2).

4. **Class weights.** Minimising the weighted log-loss with $w_k = n/(3n_k)$ targets $P_k^w \propto w_k P_k$, so $\log[P_k^w/P_0^w] = \log[P_k/P_0] + \log(w_k/w_0)$ (main eq. 3). Intercepts move, contrasts do not. For HCC, $\log(n_0/n_1) = \log(13{,}092/771) = 2.83$.

5. **Binary against ternary.** The binary target pools classes 0 and 2: $\operatorname{logit} P(Y = 1 \mid x) = L_H - \log(1 + e^{L_D})$, with $L_k = \log(P_k/P_0)$. Hence

$$\frac{\partial}{\partial x_j} \operatorname{logit} P(Y = 1 \mid x) = \frac{\partial g_j^{(1)}}{\partial x_j} - q(x) \frac{\partial g_j^{(2)}}{\partial x_j}, \qquad q = \frac{P_2}{P_0 + P_2}.$$

An additive binary model approximates $h_j' \approx g_j^{(1)\prime} - \mathrm{E}[q \mid x_j]\, g_j^{(2)\prime}$ (main eq. 4). $q$ is estimated from the ternary model after removing the class-weight shift of item 4 (Supplementary Fig. 7).

6. **Absolute risk from the deployment model.** Risk is governed by $F(\tau \mid x, d) = 1 - [1 - F_0(\tau)]^{\exp(\gamma_1 L_H + \gamma_2 L_D + \gamma_3 \log d)}$, where $d$ represent years since the cirrhosis record (main eq. 5). Fine–Gray partial likelihood on stacked laboratory visits (one per patient per 6 months, 2020–2024) with out-of-fold $L_H$ and $L_D$: a landmark supermodel (1). $F_0$ is the baseline cumulative incidence at $L_H = L_D = 0$, $d = 1$. Medians over 20 repeats (Supplementary Table 6). Substituting item 3: $\gamma_1 L_H + \gamma_2 L_D = \text{const} + \sum_j s_j(x_j)$, with $s_j = \gamma_1 g_j^{(1)} + \gamma_2 g_j^{(2)}$, and $\exp(s_j)$ is a subdistribution hazard ratio (main eq. 6).

7. **Discrimination.** AUC($\tau$), the cumulative/dynamic time-dependent AUC for cause 1 at $\tau$ (distinct from the time-fixed AUROC of the cross-validation, where every 12-month class is known): cases $T \le \tau$ with cause 1; controls everyone else, including cause 2 by $\tau$; inverse probability of censoring weights (2). $\Delta C$: Wolbers' cause-specific concordance with follow-up censored at 36 months, influence-function CI (3).

8. **Sensitivity at equal load.** For the top fraction $q$ of a marker, $\text{Sens}_q(h) = P(\text{flagged} \mid \text{HCC by } h) = q\,\hat{F}_{\text{flag}}(h)/\hat{F}_{\text{all}}(h)$, by Bayes' rule with Aalen–Johansen $\hat{F}$ (4).

9. **Accuracy.** IPCW Brier score BS($\tau$) and IPA = $1 - \text{BS}_{\text{model}}/\text{BS}_{\text{null}}$, where the null model is the covariate-free Aalen–Johansen estimate (5). O/E = mean predicted / observed Aalen–Johansen risk. Calibration slope: the Fine–Gray coefficient of the linear predictor (6).

10. **Cross-validation inference.** For fold differences $d_i$, $i = 1, \ldots, 100$: corrected $t = \bar{d}/\sqrt{(1/100 + n_{\text{test}}/n_{\text{train}})\, s^2}$, with $n_{\text{test}}/n_{\text{train}} = 1/4$ (7); Bayesian correlated t-test, posterior Student-t with correlation $\rho = n_{\text{test}}/n$, giving $P(d < -0.01)$, $P(|d| \leq 0.01)$ and $P(d > 0.01)$ (8); Bonferroni over the three comparisons.

### Supplementary Figures

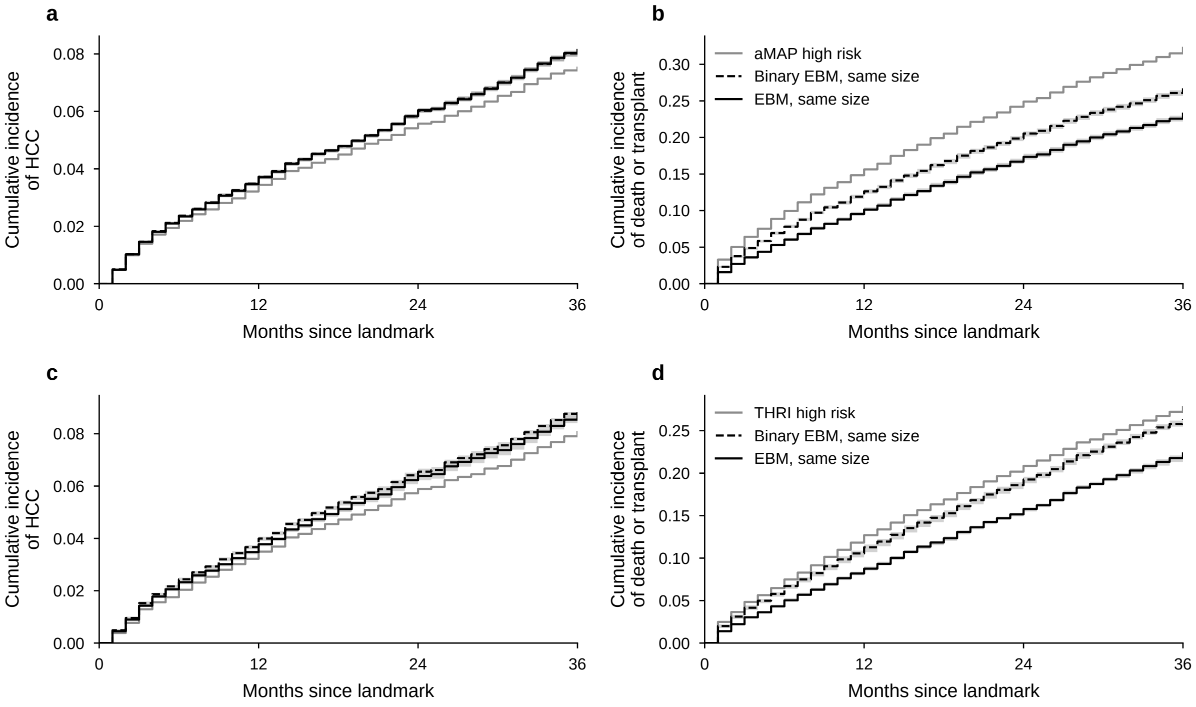


Supplementary Fig. 1. **The flagged groups over time.** Aalen–Johansen cumulative incidence of HCC (left) and of death or transplantation (right) over 36 months in three groups of the same size: the score's high-risk category (grey), the binary EBM's top-ranked group (dashed) and the model's top-ranked group (black). aMAP (a, b) and THRI (c, d) landmark sets. The excess of deaths in the score's group appears from the first months.

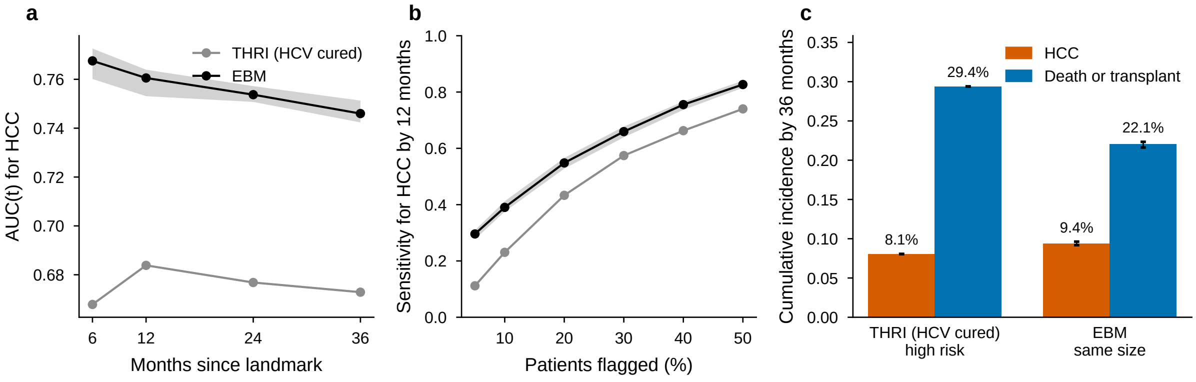


Supplementary Fig. 2. **THRI with hepatitis C taken as cured.** As Fig. 2 (a–c), with every patient with hepatitis C scored as cured (zero etiology points). Sustained virological response is not recorded, so the two versions bound each patient's THRI. This version's AUC(12 mo) is 0.684; ΔAUC(12 mo) +0.077; washout ΔC +0.056.

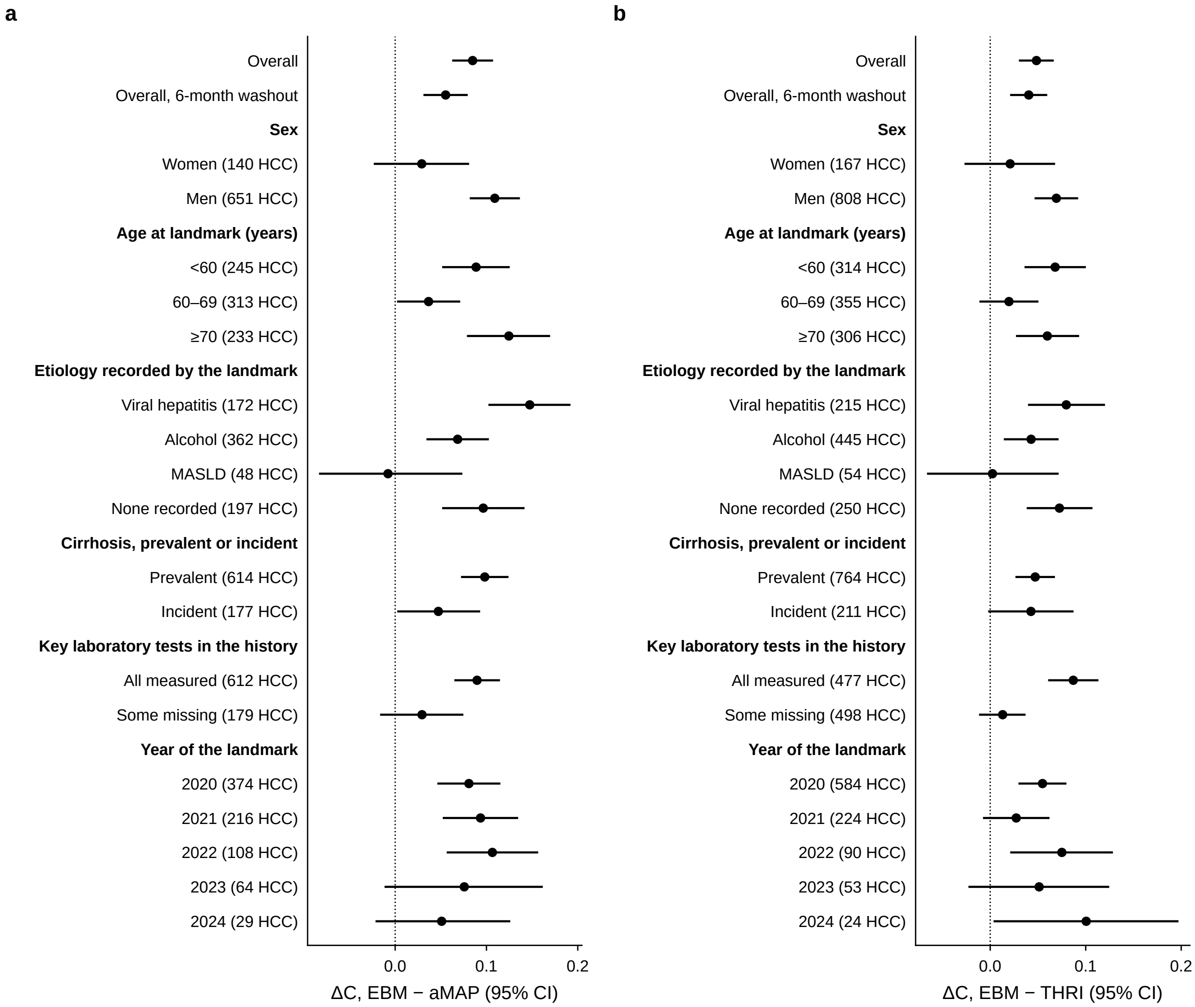


Supplementary Fig. 3. **Subgroups.** Difference in cause-specific concordance to 36 months, model minus score (95% CI), overall, after the washout, and within subgroups at the landmark (sex, age, etiology recorded by the landmark, prevalent or incident cirrhosis, whether AFP, platelets, albumin and bilirubin were all measured in the 12-month history, and calendar year of the landmark). Medians over 20 repeats of the estimate and of its CI bounds. The number of HCC cases is in parentheses. Subgroups with fewer than 10 HCC cases are not shown. The advantage holds in men, the elderly, viral hepatitis and every landmark year. It is not significant in women (140 and 167 HCC), absent in MASLD (48 and 54 HCC), and confined to patients with all four key tests measured.

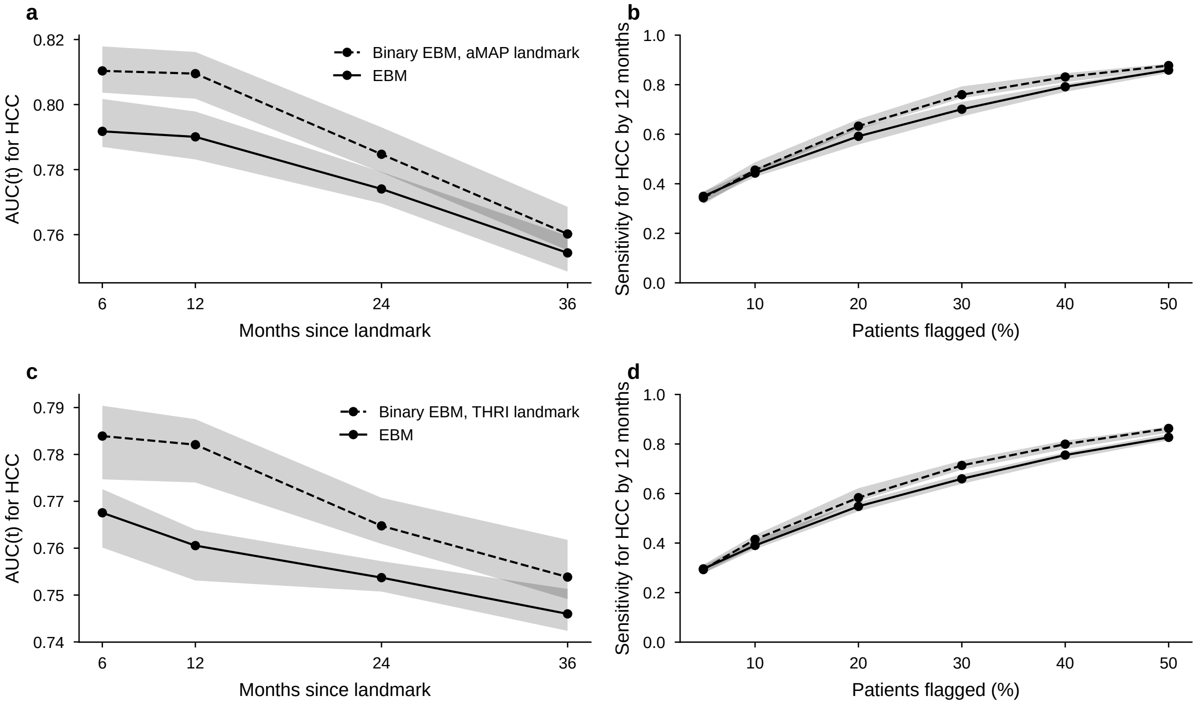


Supplementary Fig. 4. **The three-class model against the binary model at the landmarks.** AUC($t$) (time-dependent, IPCW) and sensitivity at equal load, as Fig. 2 a, b, d and e, with the binary EBM (dashed) in the place of the score. Same patients and folds. The binary model ranks 12-month HCC slightly better (ΔAUC(12 mo) -0.019 and -0.021). Over 36 months the difference shrinks (ΔC -0.006 and -0.007; Supplementary Table 4).

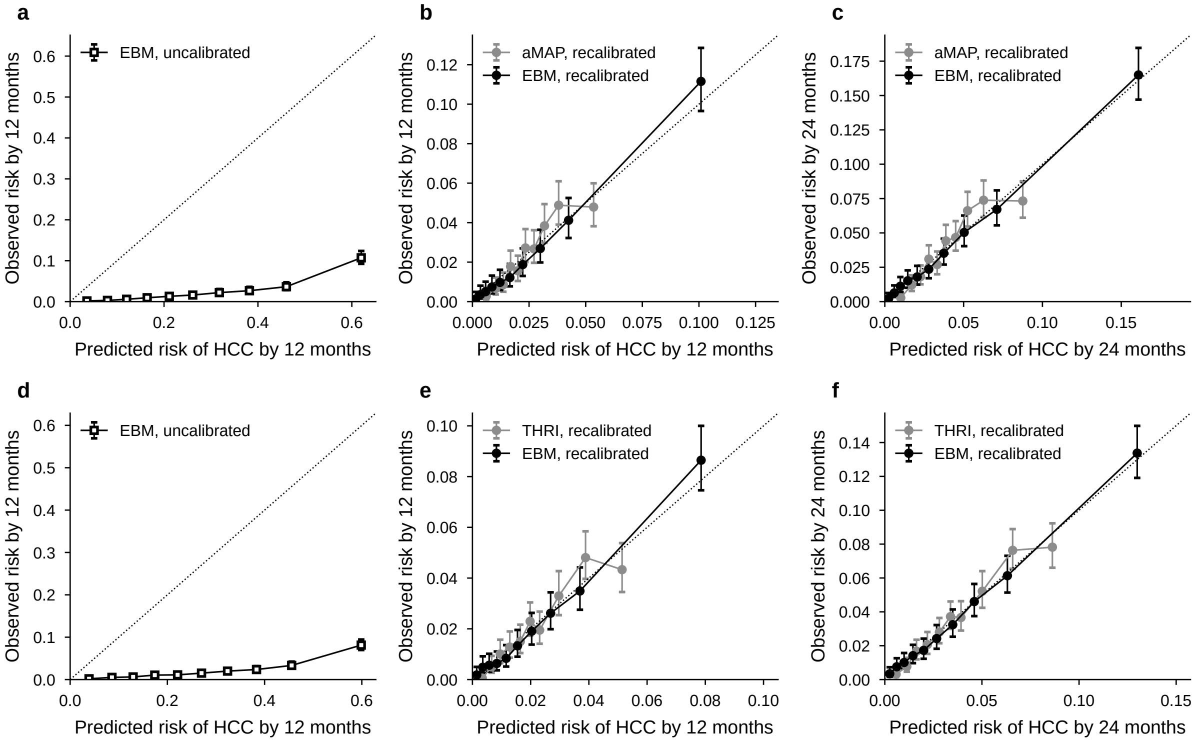


Supplementary Fig. 5. **Calibration at the score landmarks.** Deciles of predicted 12-month (a, b, d, e) and 24-month (c, f) risk against the Aalen–Johansen observed risk. a, d: the raw model output, which, with balanced class weights, ranks patients but overstates absolute risk (Methods, eq. 3). b, c, e, f: the model and the score after the same cross-fitted Fine–Gray recalibration. IPA at 12 months: model 9.5% and 6.3%, scores 0.8% and 1.0%.

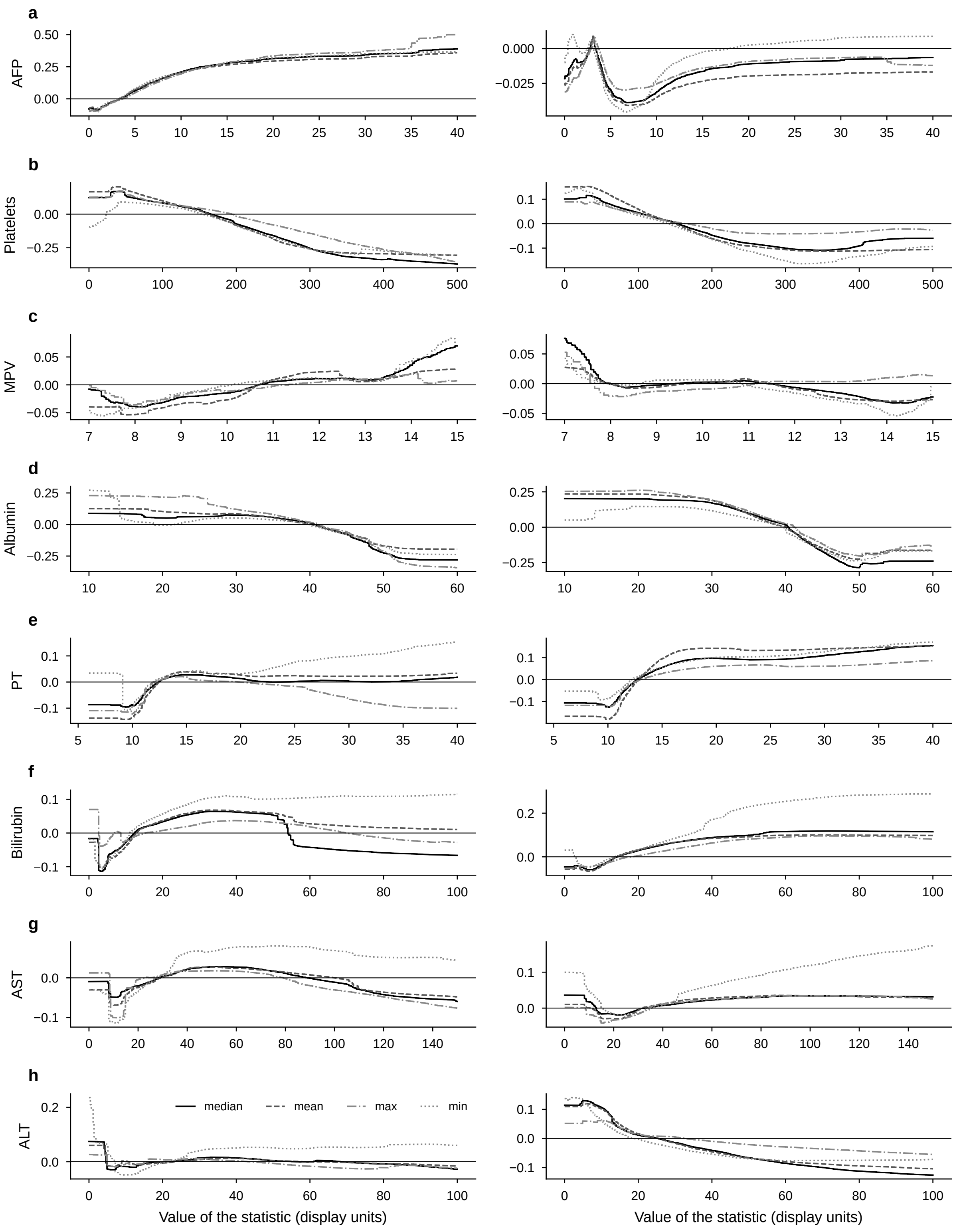


Supplementary Fig. 6. **The five statistics of each laboratory test.** Contrasts against event-free for the mean, median, maximum and minimum of each test over the 12-month window (left, HCC; right, death or transplant); the SD, on its own scale, is not drawn. Correlated statistics share the signal, so a test's effect is the sum of its five terms; hence Fig. 3a sums them, and Fig. 3c–j show the median. AFP's SD adds an HCC-only signal (+0.25 at 2 µg/L, +0.54 at 4, +0.66 at 7). The bilirubin step at 51–55 µmol/L sits in the median term; the mean and maximum decline smoothly. The minimum of bilirubin and of prothrombin time carries part of the death signal.

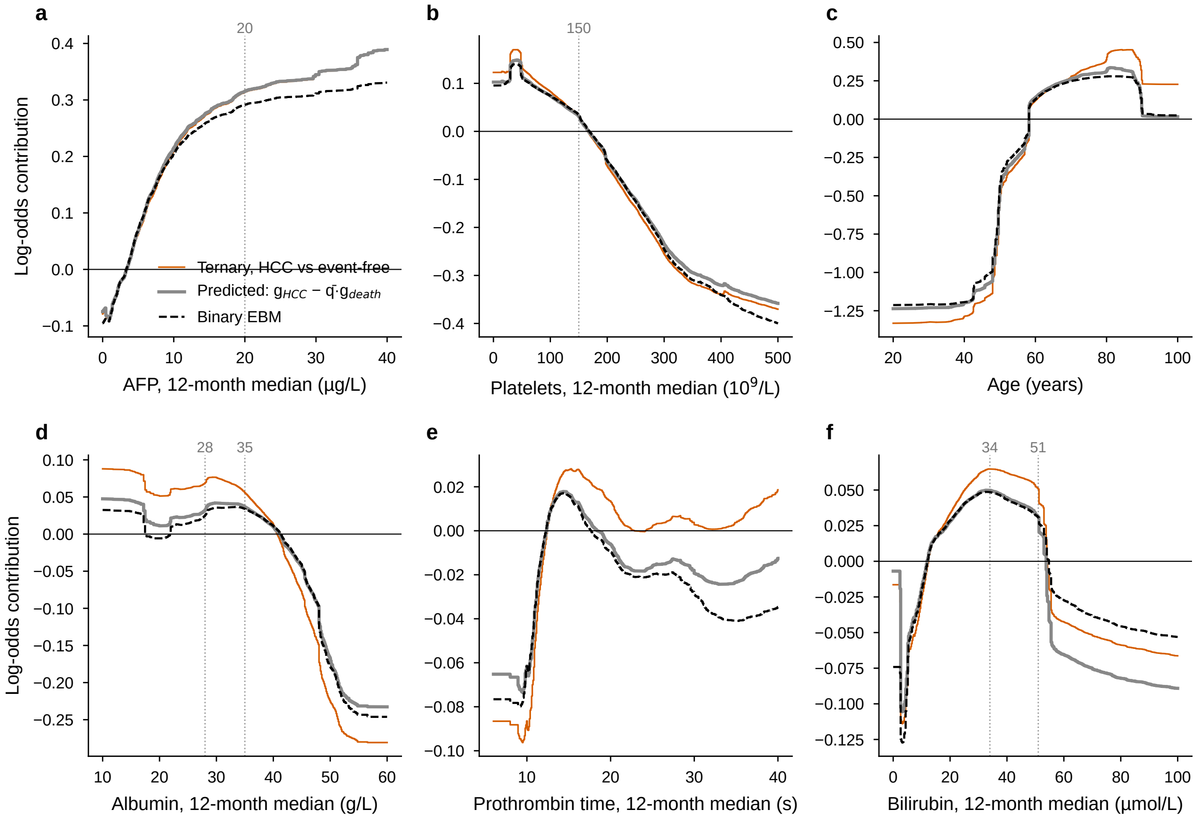


Supplementary Fig. 7. **What the binary model's curves are made of.** The binary EBM's shape function (dashed) against its first-order prediction from the three-class model, $g_{\mathrm{HCC}} - \bar{q}\, g_{\mathrm{death}}$ (grey; Methods, eq. 4), and against the HCC contrast alone (vermilion). $\bar{q} = 0.20$ is the share of deaths among patients without HCC (a local $q$, after removing the class-weight shift, fits no better). Density-weighted RMSE of the binary curve from the HCC contrast alone and from the prediction: albumin 0.030 and 0.004; prothrombin time 0.014 and 0.002; age 0.080 and 0.029; platelets 0.012 and 0.005; AFP 0.004 and 0.003; bilirubin 0.009 and 0.009. So the binary curve is the HCC curve with the death signal folded in at a fifth of its weight: the two models look alike wherever the death signal is small, the binary curve bends down where it is large (low albumin, long prothrombin time, age over 85), and it never shows the death signal itself.

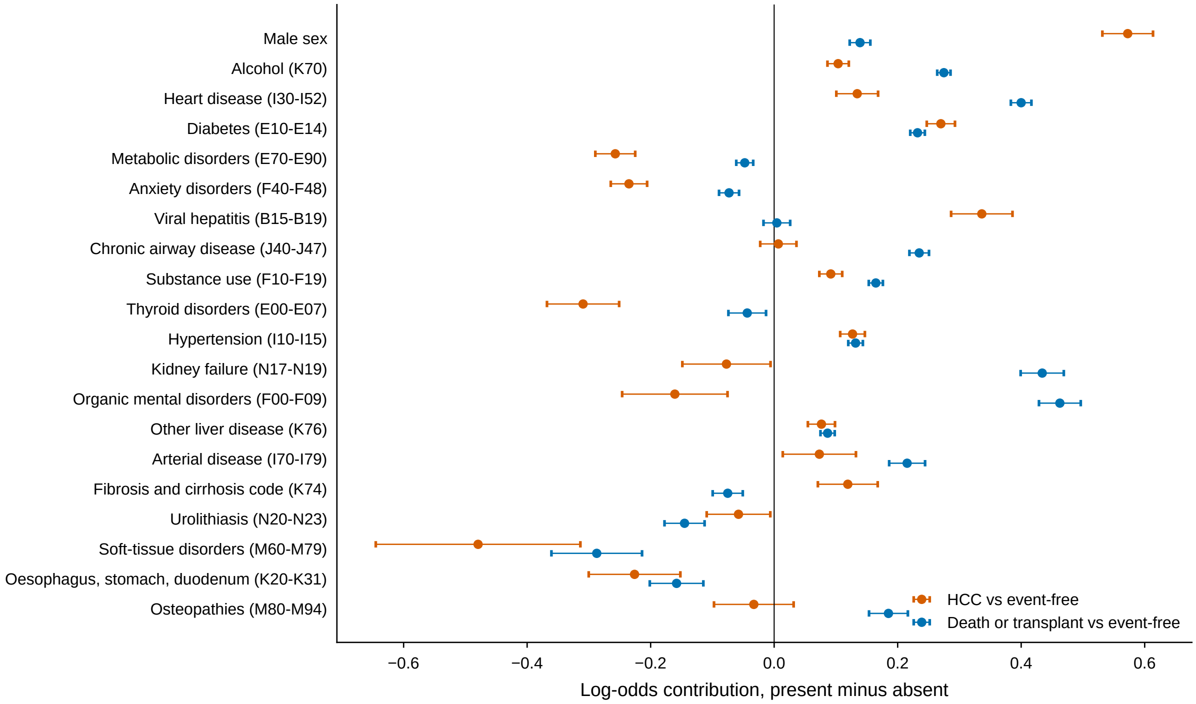


Supplementary Fig. 8. **Binary features.** The 20 most important binary features, sex and diagnosis groups (ICD-10-CM blocks of the chronic-condition registry), each as the contrast when present minus when absent, fold mean ± s.d. All continuous variables are in Fig. 3.

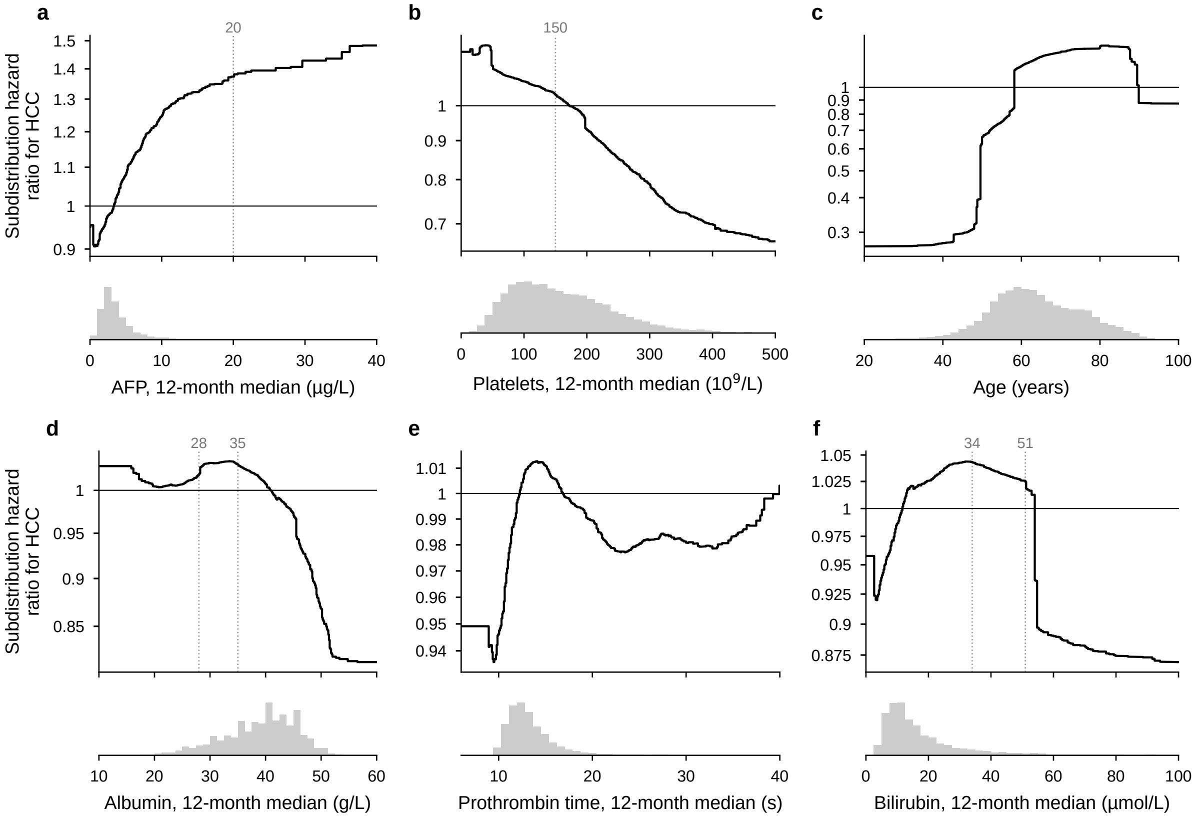


Supplementary Fig. 9. **The deployment model on the absolute-risk scale.** For each of Fig. 3's variables, the subdistribution hazard ratio for HCC contributed by its value, $\exp(\gamma_1 g_{\text{HCC}} + \gamma_2 g_{\text{death}})$ (Methods, eq. 6), from the deployment model file, centred over the cohort. Because $\gamma_2 < 0$, a value that raises the odds of death lowers the incidence of HCC: the competing risk, now in the risk itself.

### Supplementary Tables

Supplementary Table 1. **Landmark sets.** Patients with the score computable and follow-up events, those never eligible when measured, the landmark set, and how many of them also had a prediction date in the development set (the same population, seen at another time zero).

| Score | With the score computable and events | Never eligible when measured | Landmark set | Also in the development set |
|---|---|---|---|---|
| aMAP | 18,045 | 3,117 | 14,928 | 13,048 |
| THRI | 20,770 | 1,922 | 18,848 | 16,013 |
| THRI, HCV cured | 20,770 | 1,922 | 18,848 | 16,013 |

Supplementary Table 2. **Full metric profile of every model, by class.** Mean ± s.d. over the 100 test folds; threshold-dependent metrics at the class thresholds set on the validation splits (HCC: 10% false-positive rate).

| Metric | Reference | Binary | With interactions | Full record |
|---|---|---|---|---|
| **HCC** | | | | |
| AUROC | 0.734 ± 0.018 | 0.741 ± 0.017 | 0.732 ± 0.017 | 0.710 ± 0.020 |
| AUPRC | 0.148 ± 0.021 | 0.146 ± 0.020 | 0.144 ± 0.022 | 0.130 ± 0.018 |
| Brier score | 0.111 ± 0.002 | 0.197 ± 0.003 | 0.108 ± 0.002 | 0.106 ± 0.002 |
| Log loss | 0.372 ± 0.005 | 0.569 ± 0.008 | 0.360 ± 0.006 | 0.368 ± 0.005 |
| Sensitivity | 0.310 ± 0.040 | 0.319 ± 0.039 | 0.306 ± 0.037 | 0.296 ± 0.037 |
| Specificity | 0.905 ± 0.009 | 0.905 ± 0.008 | 0.905 ± 0.008 | 0.905 ± 0.009 |
| PPV | 0.134 ± 0.015 | 0.137 ± 0.014 | 0.132 ± 0.014 | 0.128 ± 0.015 |
| NPV | 0.965 ± 0.002 | 0.966 ± 0.002 | 0.965 ± 0.002 | 0.965 ± 0.002 |
| F1 | 0.187 ± 0.020 | 0.191 ± 0.019 | 0.184 ± 0.020 | 0.178 ± 0.020 |
| Balanced accuracy | 0.608 ± 0.019 | 0.612 ± 0.018 | 0.606 ± 0.018 | 0.600 ± 0.017 |
| MCC | 0.146 ± 0.024 | 0.152 ± 0.023 | 0.143 ± 0.023 | 0.136 ± 0.023 |
| LR+ | 3.295 ± 0.412 | 3.374 ± 0.401 | 3.242 ± 0.409 | 3.131 ± 0.419 |
| LR- | 0.762 ± 0.041 | 0.752 ± 0.041 | 0.767 ± 0.040 | 0.778 ± 0.039 |
| **Death or transplant without HCC** | | | | |
| AUROC | 0.759 ± 0.008 | – | 0.764 ± 0.008 | 0.773 ± 0.008 |
| AUPRC | 0.412 ± 0.014 | – | 0.420 ± 0.014 | 0.429 ± 0.015 |
| Brier score | 0.150 ± 0.002 | – | 0.150 ± 0.002 | 0.146 ± 0.002 |
| Log loss | 0.465 ± 0.005 | – | 0.462 ± 0.006 | 0.459 ± 0.004 |
| Sensitivity | 0.370 ± 0.019 | – | 0.404 ± 0.019 | 0.321 ± 0.019 |
| Specificity | 0.890 ± 0.008 | – | 0.877 ± 0.009 | 0.917 ± 0.007 |
| PPV | 0.447 ± 0.018 | – | 0.440 ± 0.017 | 0.481 ± 0.019 |
| NPV | 0.855 ± 0.004 | – | 0.860 ± 0.004 | 0.850 ± 0.003 |
| F1 | 0.405 ± 0.016 | – | 0.421 ± 0.014 | 0.385 ± 0.017 |
| Balanced accuracy | 0.630 ± 0.009 | – | 0.640 ± 0.009 | 0.619 ± 0.009 |
| MCC | 0.280 ± 0.018 | – | 0.290 ± 0.017 | 0.281 ± 0.018 |
| LR+ | 3.384 ± 0.247 | – | 3.293 ± 0.230 | 3.894 ± 0.309 |
| LR- | 0.707 ± 0.020 | – | 0.680 ± 0.020 | 0.740 ± 0.020 |
| **Event-free** | | | | |
| AUROC | 0.752 ± 0.008 | – | 0.756 ± 0.008 | 0.754 ± 0.008 |
| AUPRC | 0.904 ± 0.004 | – | 0.907 ± 0.004 | 0.900 ± 0.005 |
| Brier score | 0.283 ± 0.003 | – | 0.274 ± 0.004 | 0.277 ± 0.003 |
| Log loss | 0.776 ± 0.008 | – | 0.758 ± 0.009 | 0.757 ± 0.007 |

| Metric | Reference | Binary | With interactions | Full record |
|---|---|---|---|---|
| Sensitivity | 0.388 ± 0.011 | – | 0.429 ± 0.013 | 0.372 ± 0.011 |
| Specificity | 0.910 ± 0.012 | – | 0.891 ± 0.013 | 0.907 ± 0.011 |
| PPV | 0.932 ± 0.008 | – | 0.927 ± 0.007 | 0.928 ± 0.007 |
| NPV | 0.317 ± 0.004 | – | 0.328 ± 0.005 | 0.311 ± 0.004 |
| F1 | 0.548 ± 0.011 | – | 0.587 ± 0.012 | 0.531 ± 0.011 |
| Balanced accuracy | 0.649 ± 0.006 | – | 0.660 ± 0.007 | 0.640 ± 0.006 |
| MCC | 0.273 ± 0.011 | – | 0.286 ± 0.011 | 0.258 ± 0.011 |
| LR+ | 4.370 ± 0.539 | – | 4.005 ± 0.439 | 4.071 ± 0.488 |
| LR- | 0.672 ± 0.012 | – | 0.640 ± 0.013 | 0.692 ± 0.012 |

Supplementary Table 3. **Comparison with the scores within subgroups.** Key laboratory tests: AFP, platelets, albumin and bilirubin, all measured or at least one missing in the 12-month history window (a missing test enters the model as the training median). Year: calendar year of the landmark; AUC(12 mo) is not estimable for 2024, with less than 12 months of follow-up. AUC(12 mo): time-dependent AUC (IPCW). O/E 12 mo: observed/expected 12-month risk under the visit-based recalibration. Medians over 20 repeats; not computed with fewer than 10 HCC cases.

| Score | Subgroup | Level | Patients | HCC | AUC(12 mo), score | AUC(12 mo), EBM | ΔC to 36 mo (95% CI) | O/E 12 mo |
|---|---|---|---|---|---|---|---|---|
| aMAP | Sex | Female | 5,225 | 140 | 0.735 | 0.795 | +0.029 (-0.022, +0.080) | 0.87 |
| aMAP | Sex | Male | 9,703 | 651 | 0.638 | 0.761 | +0.109 (+0.083, +0.135) | 1.19 |
| aMAP | Age, years | <60 | 6,063 | 245 | 0.729 | 0.830 | +0.089 (+0.053, +0.124) | 1.08 |
| aMAP | Age, years | 60–69 | 4,655 | 313 | 0.696 | 0.763 | +0.037 (+0.003, +0.070) | 1.19 |
| aMAP | Age, years | ≥ 70 | 4,210 | 233 | 0.628 | 0.758 | +0.125 (+0.080, +0.169) | 1.09 |
| aMAP | Etiology | Viral hepatitis | 2,301 | 172 | 0.622 | 0.805 | +0.147 (+0.103, +0.191) | 1.22 |
| aMAP | Etiology | Alcohol | 6,222 | 362 | 0.684 | 0.742 | +0.068 (+0.035, +0.101) | 1.12 |
| aMAP | Etiology | MASLD | 1,529 | 48 | 0.774 | 0.807 | -0.008 (-0.082, +0.072) | 0.88 |
| aMAP | Etiology | Other | 1,370 | 12 | – | – | – | 0.33 |
| aMAP | Etiology | None recorded | 3,506 | 197 | 0.656 | 0.798 | +0.096 (+0.053, +0.141) | 1.21 |
| aMAP | Cirrhosis | Prevalent | 8,869 | 614 | 0.665 | 0.782 | +0.098 (+0.073, +0.123) | 1.10 |
| aMAP | Cirrhosis | Incident | 6,059 | 177 | 0.731 | 0.808 | +0.047 (+0.003, +0.092) | 1.14 |
| aMAP | Key laboratory tests in the history | All measured | 9,339 | 612 | 0.683 | 0.793 | +0.090 (+0.066, +0.114) | 1.22 |

| Score | Subgroup | Level | Patients | HCC | AUC(12 mo), score | AUC(12 mo), EBM | ΔC to 36 mo (95% CI) | O/E 12 mo |
|---|---|---|---|---|---|---|---|---|
| aMAP | Key laboratory tests in the history | Some missing | 5,589 | 179 | 0.707 | 0.736 | +0.029 (-0.015, +0.074) | 0.84 |
| aMAP | Year of the landmark | 2020 | 4,908 | 374 | 0.659 | 0.748 | +0.081 (+0.047, +0.114) | 0.99 |
| aMAP | Year of the landmark | 2021 | 3,325 | 216 | 0.675 | 0.791 | +0.094 (+0.053, +0.134) | 1.23 |
| aMAP | Year of the landmark | 2022 | 2,557 | 108 | 0.677 | 0.817 | +0.106 (+0.058, +0.155) | 0.96 |
| aMAP | Year of the landmark | 2023 | 2,132 | 64 | 0.752 | 0.840 | +0.076 (-0.010, +0.161) | 1.36 |
| aMAP | Year of the landmark | 2024 | 2,006 | 29 | – | – | +0.051 (-0.020, +0.125) | – |
| THRI | Sex | Female | 6,503 | 167 | 0.743 | 0.785 | +0.021 (-0.026, +0.067) | 0.72 |
| THRI | Sex | Male | 12,345 | 808 | 0.636 | 0.723 | +0.069 (+0.048, +0.091) | 1.13 |
| THRI | Age, years | <60 | 7,780 | 314 | 0.701 | 0.792 | +0.068 (+0.037, +0.099) | 1.11 |
| THRI | Age, years | 60–69 | 5,661 | 355 | 0.712 | 0.721 | +0.020 (-0.010, +0.049) | 1.02 |
| THRI | Age, years | ≥ 70 | 5,407 | 306 | 0.678 | 0.750 | +0.060 (+0.028, +0.092) | 1.02 |
| THRI | Etiology | Viral hepatitis | 2,960 | 215 | 0.612 | 0.740 | +0.080 (+0.041, +0.119) | 1.12 |
| THRI | Etiology | Alcohol | 7,647 | 445 | 0.674 | 0.721 | +0.043 (+0.015, +0.071) | 1.13 |
| THRI | Etiology | MASLD | 1,896 | 54 | 0.739 | 0.840 | +0.002 (-0.065, +0.071) | 0.66 |
| THRI | Etiology | Other | 1,626 | 11 | – | – | – | 0.20 |
| THRI | Etiology | None recorded | 4,719 | 250 | 0.684 | 0.770 | +0.072 (+0.039, +0.106) | 1.03 |
| THRI | Cirrhosis | Prevalent | 10,949 | 764 | 0.676 | 0.735 | +0.047 (+0.028, +0.067) | 1.01 |
| THRI | Cirrhosis | Incident | 7,899 | 211 | 0.757 | 0.805 | +0.043 (-0.001, +0.086) | 1.09 |
| THRI | Key laboratory tests in the history | All measured | 7,923 | 477 | 0.694 | 0.789 | +0.087 (+0.062, +0.112) | 1.07 |

| Score | Subgroup | Level | Patients | HCC | AUC(12 mo), score | AUC(12 mo), EBM | ΔC to 36 mo (95% CI) | O/E 12 mo |
|---|---|---|---|---|---|---|---|---|
| THRI | Key laboratory tests in the history | Some missing | 10,925 | 498 | 0.705 | 0.726 | +0.013 (-0.011, +0.036) | 1.02 |
| THRI | Year of the landmark | 2020 | 8,261 | 584 | 0.685 | 0.742 | +0.055 (+0.031, +0.079) | 0.96 |
| THRI | Year of the landmark | 2021 | 3,506 | 224 | 0.704 | 0.747 | +0.027 (-0.006, +0.061) | 1.21 |
| THRI | Year of the landmark | 2022 | 2,684 | 90 | 0.711 | 0.794 | +0.075 (+0.022, +0.127) | 0.82 |
| THRI | Year of the landmark | 2023 | 2,255 | 53 | 0.745 | 0.792 | +0.051 (-0.022, +0.123) | 1.23 |
| THRI | Year of the landmark | 2024 | 2,142 | 24 | – | – | +0.101 (+0.005, +0.196) | – |

Supplementary Table 4. **The three-class model against the binary EBM at the score landmarks.** Same patients, folds and repeats; differences are ternary minus binary.

| | aMAP landmark | aMAP landmark, 6-month washout | THRI landmark | THRI landmark, 6-month washout |
|---|---|---|---|---|
| Patients | 14,928 | 12,803 | 18,848 | 16,547 |
| HCC | 791 | 567 | 975 | 743 |
| Death/transplant | 3,492 | 2,456 | 4,547 | 3,441 |
| AUC(6 mo), binary EBM | 0.810 | 0.807 | 0.784 | 0.780 |
| AUC(6 mo), ternary EBM | 0.792 | 0.783 | 0.768 | 0.749 |
| ΔAUC(6 mo) (95% CI) | -0.019 (-0.031, -0.005) | -0.026 (-0.043, -0.008) | -0.016 (-0.027, -0.005) | -0.033 (-0.051, -0.016) |
| AUC(12 mo), binary EBM | 0.810 | 0.784 | 0.782 | 0.772 |
| AUC(12 mo), ternary EBM | 0.790 | 0.762 | 0.761 | 0.748 |
| ΔAUC(12 mo) (95% CI) | -0.019 (-0.029, -0.008) | -0.021 (-0.036, -0.009) | -0.021 (-0.032, -0.011) | -0.024 (-0.034, -0.011) |
| AUC(24 mo), binary EBM | 0.785 | 0.765 | 0.765 | 0.756 |
| AUC(24 mo), ternary EBM | 0.774 | 0.752 | 0.754 | 0.746 |
| ΔAUC(24 mo) (95% CI) | -0.011 (-0.018, -0.003) | -0.013 (-0.025, -0.004) | -0.011 (-0.019, -0.004) | -0.011 (-0.019, -0.002) |
| AUC(36 mo), binary EBM | 0.760 | 0.740 | 0.754 | 0.744 |

| | aMAP landmark | aMAP landmark, 6-month washout | THRI landmark | THRI landmark, 6-month washout |
|---|---|---|---|---|
| AUC(36 mo), ternary EBM | 0.754 | 0.734 | 0.746 | 0.738 |
| ΔAUC(36 mo) (95% CI) | -0.007 (-0.014, +0.000) | -0.006 (-0.014, +0.002) | -0.008 (-0.014, -0.001) | -0.006 (-0.013, +0.001) |
| ΔC to 36 mo (95% CI) | -0.006 (-0.013, +0.001) | -0.005 (-0.012, +0.003) | -0.007 (-0.014, -0.001) | -0.006 (-0.013, +0.001) |

Supplementary Table 5. **Comparison with THRI with hepatitis C scored as cured.**

| | THRI (HCV cured) | THRI (HCV cured), 6-month washout |
|---|---|---|
| Patients | 18,848 | 16,547 |
| HCC | 975 | 743 |
| Death/transplant | 4,547 | 3,441 |
| AUC(6 mo), score | 0.668 | 0.714 |
| AUC(6 mo), EBM | 0.768 | 0.749 |
| ΔAUC(6 mo) (95% CI) | +0.100 (+0.061, +0.136) | +0.036 (-0.003, +0.077) |
| AUC(12 mo), score | 0.684 | 0.712 |
| AUC(12 mo), EBM | 0.761 | 0.748 |
| ΔAUC(12 mo) (95% CI) | +0.077 (+0.048, +0.106) | +0.037 (+0.009, +0.065) |
| AUC(24 mo), score | 0.677 | 0.689 |
| AUC(24 mo), EBM | 0.754 | 0.746 |
| ΔAUC(24 mo) (95% CI) | +0.077 (+0.058, +0.097) | +0.057 (+0.036, +0.081) |
| AUC(36 mo), score | 0.673 | 0.684 |
| AUC(36 mo), EBM | 0.746 | 0.738 |
| ΔAUC(36 mo) (95% CI) | +0.073 (+0.056, +0.094) | +0.055 (+0.034, +0.076) |
| ΔC to 36 mo (95% CI) | +0.071 (+0.052, +0.091) | +0.056 (+0.036, +0.076) |

Supplementary Table 6. **Visit-based recalibration and size of the deployment model.** Coefficients of the Fine–Gray recalibration (eq. 5) and baseline cumulative incidence, medians and ranges over 20 repeats, and the number of stored values in the deployment model file.

| Parameter | Median over 20 repeats | Range |
|---|---|---|
| $\gamma_1$, log-odds of HCC vs event-free | 1.053 | 1.012 to 1.107 |
| $\gamma_2$, log-odds of death or transplant vs event-free | -0.283 | -0.322 to -0.261 |
| $\gamma_3$, log years since the cirrhosis record | 0.106 | 0.099 to 0.113 |
| $F_0$(6 months) | 0.0099 | 0.0096 to 0.0101 |
| $F_0$(12 months) | 0.0170 | 0.0164 to 0.0173 |
| $F_0$(24 months) | 0.0296 | 0.0287 to 0.0301 |
| $F_0$(36 months) | 0.0418 | 0.0405 to 0.0425 |
| Model file: terms | 110 | |
| Model file: features | 110 | |
| Model file: bin scores | 92,988 | |
| Model file: intercepts | 3 | |
| Model file: recalibration coefficients | 3 | |
| Model file: baseline cif values | 4 | |

Supplementary Table 7. **The 110 features of the model,** derived automatically from age, sex, coded chronic diagnoses and eight laboratory tests. Diagnosis groups are ICD-10-CM blocks (liver diseases K70–K77 at category level); "unclassified" collects chronic-registry records without a code.

| Group | Features | Content |
|---|---|---|
| Demographic | 2 | Age (at the index date in development, at time zero in evaluation and deployment); male sex |
| Diagnosis groups (chronic-condition registry) | 68 | B15-B19, B20-B24, C00-C75, D60-D64, D65-D69, E00-E07, E10-E14, E65-E68, E70-E90, F00-F09, F10-F19, F20-F29, F30-F39, F40-F48, F50-F59, F60-F69, F70-F79, F80-F89, F90-F98, F99-F99, G10-G14, G20-G26, G30-G32, G35-G37, G40-G47, G60-G64, G70-G73, G80-G83, G90-G99, H30-H36, H40-H42, I05-I09, I10-I15, I20-I25, I26-I28, I30-I52, I60-I69, I70-I79, I95-I99, J40-J47, J60-J70, K20-K31, K50-K52, K55-K63, K70, K71, K73, K74, K75, K76, K90-K93, L20-L30, L40-L45, M00-M25, M30-M36, M40-M54, M60-M79, M80-M94, N17-N19, N20-N23, Q00-Q07, Q20-Q28, R40-R46, T80-T88, Z20-Z29, Z40-Z54, Z80-Z99, unclassified |
| Laboratory tests (mean, median, SD, maximum, minimum over 12 months) | 40 | Mean platelet volume (OMOP 3001123); Total bilirubin (OMOP 3006140); ALT (OMOP 3006923); AST (OMOP 3013721); Albumin (OMOP 3024561); Platelets (OMOP 3024929); Prothrombin time (OMOP 3034426); Alpha-fetoprotein (OMOP 3039783) |

Supplementary Table 8. **Configuration of the reference model.** Cross-validation, class weights, preprocessing and every constructor argument of the Explainable Boosting Machine, defaults included, as used for all models (the variants change only the outcome, the interactions or the features).

| Setting | Value | Note |
|---|---|---|
| **Cross-validation** | | |
| repeats x folds | 20 x 5 | stratified by the three-class outcome; seed random_state + repeat |
| validation fraction | 0.2 | of each training part, for the operating threshold |
| operating threshold | FPR 0.1 | HCC, on the validation split |
| class weights w_k | n / (3 n_k) | balanced, from each training subset |
| **Preprocessing (scikit-learn)** | | |
| age | StandardScaler | |
| sex, diagnosis groups | binary flags | passed through |
| laboratory statistics | RobustScaler, then SimpleImputer (median) | a missing value takes the training median |
| **Explainable Boosting Machine (interpret 0.6.12)** | | |
| `cat_smooth` | `10.0` | |
| `cyclic_progress` | `False` | |
| `early_stopping_rounds` | `100` | |
| `early_stopping_tolerance` | `1e-05` | |
| `exclude` | `None` | |
| `feature_names` | `None` | |
| `feature_types` | `None` | |
| `gain_scale` | `5.0` | |
| `greedy_ratio` | `10.0` | |
| `inner_bags` | `0` | |
| `interaction_smoothing_rounds` | `75` | |
| `interactions` | `0` | set in params.yaml (default '3x') |

| Setting | Value | Note |
|---|---|---|
| `learning_rate` | `0.015` | |
| `max_bins` | `1024` | |
| `max_delta_step` | `0.0` | |
| `max_interaction_bins` | `64` | |
| `max_leaves` | `2` | |
| `max_rounds` | `50000` | |
| `min_cat_samples` | `10` | |
| `min_hessian` | `0.0001` | |
| `min_samples_leaf` | `4` | |
| `missing` | `'separate'` | |
| `monotone_constraints` | `None` | |
| `n_jobs` | `-2` | 1 within each cross-validation fold (folds run in parallel) |
| `objective` | `'log_loss'` | |
| `outer_bags` | `14` | |
| `random_state` | `42` | general_params.random_state |
| `reg_alpha` | `0.0` | |
| `reg_lambda` | `0.0` | |
| `smoothing_rounds` | `75` | |
| `validation_size` | `0.15` | |

Supplementary Table 9. **THRI etiology codes.** With more than one etiology recorded by the landmark, the highest-scoring one is used. Sustained virological response is not recorded; the HCV-cured version scores hepatitis C as 0.

| THRI etiology | ICD-10-CM codes | Points |
|---|---|---:|
| HCV | B17.1, B18.2, B19.2 | 97 |
| HBV | B16, B17.0, B18.0, B18.1, B19.1 | 97 |
| Steatohepatitis | K70, K75.81, K76.0 | 54 |
| Autoimmune | K75.4, K74.3, K83.01 | 0 |
| Other | E83.11, E83.01, E88.01, E80.0, E80.1, E80.2 | 36 |
| None recorded (cryptogenic, scored as Other) | – | 36 |

Supplementary Table 10. **Internal–external validation by province.** The reference model refitted on the development patients of the other seven provinces, with the deployment model's pipeline, hyperparameters and class weights, and applied to the held-out province. Patients, HCC and AUROC: development design (12-month outcome), held out against the cross-validated predictions for the same patients (CV, median over 20 repeats); the CV models were trained on 64% of the patients, the province models on 77–94%. ΔC: difference in cause-specific concordance to 36 months, held-out-province model minus score, at each score's landmark (95% CI by influence functions). All provinces: pooled by a DerSimonian–Laird random-effects model, with $I^2$. Provinces are assigned from the patient's primary-care health district.

| Province | Patients | HCC | HCC AUROC, held out | HCC AUROC, CV | Death AUROC, held out | Death AUROC, CV | ΔC vs aMAP (95% CI) | ΔC vs THRI (95% CI) |
|---|---|---|---|---|---|---|---|---|
| Almería | 1,432 | 42 | 0.713 | 0.703 | 0.796 | 0.793 | +0.029 (-0.057 to +0.115) | +0.068 (-0.002 to +0.138) |

| Province | Patients | HCC | HCC AUROC, held out | HCC AUROC, CV | Death AUROC, held out | Death AUROC, CV | ΔC vs aMAP (95% CI) | ΔC vs THRI (95% CI) |
|---|---|---|---|---|---|---|---|---|
| Cádiz | 2,928 | 143 | 0.726 | 0.720 | 0.758 | 0.760 | +0.100 (+0.054 to +0.146) | +0.053 (+0.017 to +0.090) |
| Córdoba | 1,838 | 76 | 0.730 | 0.724 | 0.791 | 0.787 | +0.038 (-0.026 to +0.101) | +0.047 (-0.006 to +0.100) |
| Granada | 2,123 | 96 | 0.736 | 0.736 | 0.739 | 0.736 | +0.088 (+0.035 to +0.142) | +0.065 (+0.022 to +0.107) |
| Huelva | 948 | 42 | 0.753 | 0.736 | 0.748 | 0.747 | +0.111 (+0.050 to +0.172) | +0.089 (+0.037 to +0.142) |
| Jaén | 1,156 | 50 | 0.777 | 0.764 | 0.741 | 0.740 | +0.167 (+0.088 to +0.245) | +0.088 (+0.008 to +0.168) |
| Málaga | 2,735 | 115 | 0.792 | 0.789 | 0.762 | 0.764 | +0.082 (+0.029 to +0.136) | +0.053 (+0.007 to +0.099) |
| Sevilla | 4,017 | 207 | 0.718 | 0.716 | 0.752 | 0.752 | +0.100 (+0.053 to +0.146) | +0.037 (+0.001 to +0.073) |
| All provinces | 17,177 | 771 | 0.738 | 0.733 | 0.759 | 0.759 | +0.091 (+0.068 to +0.114); $I^2$ 23% | +0.057 (+0.040 to +0.073); $I^2$ 0% |

Supplementary Table 11. **The visit-based recalibration by sex.** Observed/expected risk of HCC at 6 to 36 months at the routine laboratory visits on which the recalibration was fitted, for the released recalibration (no sex term) and for the same refitted with a female indicator, both cross-fitted by fold; medians over 20 repeats. A ratio below 1 means overestimated risk. Female subdistribution hazard ratio in the refits: median 0.98 (range 0.81 to 1.11).

| Recalibration | Patients | 6 mo | 12 mo | 24 mo | 36 mo |
|---|---|---|---|---|---|
| Visit-based recalibration | All | 1.00 | 1.00 | 1.01 | 1.02 |
| Visit-based recalibration | Women | 0.83 | 0.89 | 0.97 | 1.02 |
| Visit-based recalibration | Men | 1.04 | 1.03 | 1.02 | 1.02 |
| Refitted with a female indicator | All | 1.00 | 1.00 | 1.01 | 1.02 |
| Refitted with a female indicator | Women | 0.85 | 0.91 | 0.99 | 1.04 |
| Refitted with a female indicator | Men | 1.03 | 1.02 | 1.01 | 1.01 |

Supplementary Table 12. **The visit-based recalibration at time zeros that are not laboratory visits.** Observed/expected risk of HCC at 6 to 36 months when the model is applied on each anniversary of the study start (2 January 2020 to 2024) to every patient eligible on that date (10,506 to 13,122 per date), with features from the previous 12 months and out-of-fold predictions; medians over 20 repeats. A ratio below 1 means overestimated risk. Laboratory visit: at least one laboratory result in the 6 months before the date. Overall AUC(12 mo) 0.75 and calibration slope 0.96, against 0.78 and 0.99 at laboratory visits; IPA at 12 months 3.4%. A dash: follow-up shorter than the horizon.

| Subgroup | Level | 6 mo | 12 mo | 24 mo | 36 mo |
|---|---|---|---|---|---|
| All time zeros | | 0.72 | 0.79 | 0.90 | 0.94 |
| Laboratory visit in the previous 6 months | Yes | 0.82 | 0.85 | 0.94 | 0.97 |
| Laboratory visit in the previous 6 months | No | 0.56 | 0.68 | 0.83 | 0.90 |
| Sex | Women | 0.54 | 0.63 | 0.75 | 0.81 |
| Sex | Men | 0.77 | 0.83 | 0.93 | 0.97 |

| Subgroup | Level | 6 mo | 12 mo | 24 mo | 36 mo |
| --- | --- | --- | --- | --- | --- |
| Age, years | <60 | 0.68 | 0.76 | 0.88 | 0.96 |
| Age, years | 60–69 | 0.79 | 0.81 | 0.93 | 0.98 |
| Age, years | ≥ 70 | 0.68 | 0.79 | 0.88 | 0.90 |
| Etiology | Viral hepatitis | 0.73 | 0.83 | 0.95 | 1.04 |
| Etiology | Alcohol | 0.74 | 0.79 | 0.86 | 0.87 |
| Etiology | MASLD | 0.59 | 0.59 | 0.75 | 0.87 |
| Etiology | Other | 0.22 | 0.26 | 0.34 | 0.32 |
| Etiology | None recorded | 0.80 | 0.89 | 1.04 | 1.11 |
| Cirrhosis | Prevalent | 0.72 | 0.81 | 0.90 | 0.93 |
| Cirrhosis | Incident | 0.71 | 0.69 | 0.83 | 0.90 |
| Year | 2020 | 0.73 | 0.81 | 0.91 | 0.92 |
| Year | 2021 | 0.72 | 0.84 | 0.90 | 0.91 |
| Year | 2022 | 0.64 | 0.72 | 0.80 | – |
| Year | 2023 | 0.64 | 0.71 | – | – |
| Year | 2024 | 0.86 | – | – | – |

Supplementary Table 13. **Recent HCC risk models in cirrhosis or chronic liver disease.** Each row checked against its abstract.

| Study | Population, data | Model, explanation | Competing risks | Comparators | Result |
| --- | --- | --- | --- | --- | --- |
| aMAP (9) | chronic hepatitis, mostly HBV, cohorts | points score | – | – | 5-year risk; benchmark |
| THRI (10) | cirrhosis, 2,079, tertiary | Cox, points | no (Cox) | – | external C 0.77; 10-year risk |
| THCC-RI (11) | cirrhosis under surveillance, 1,560, 7 US centres | regression index | censoring at transplant or death | aMAP | C 0.77; similar to aMAP to 2 years |
| Audureau 2020 (12) | HCV cirrhosis, biopsy-proven, 836 (CirVir) | random survival forest | yes | Fine–Gray | external C 0.70–0.71 vs 0.64 |
| Ioannou 2020 (13) | HCV cirrhosis, 48,151, VA EHR | RNN on raw records, black box | no (binary, 3 years) | logistic regression | AUROC 0.76 vs 0.69 |
| PLAN-B (14) | CHB on antivirals, 6,051 + 7,457 | gradient boosting | no | PAGE-B, CU-HCC, REACH-B | C 0.79–0.81 |
| Nam 2020 (15) | HBV cirrhosis on entecavir, 424 + 316 | deep learning | no | 6 scores incl. THRI | C 0.78 vs THRI 0.59 |
| SMART-HCC (16) | chronic liver disease, 5,155 + 5,116 | ML with liver stiffness | no | aMAP, THRI, 7 HBV scores | C 0.89–0.91 (needs elastography) |
| Nahon 2026 (17) | compensated cirrhosis (cured HCV 73%, controlled HBV 14%, alcohol or metabolic 13%), 4,867, three prospective French cohorts under semiannual surveillance | decision tree, random survival forest, DeepHit | yes (death; transplant censored) | aMAP, FASTRAK | validation time-dependent AUC at 1, 3, 5 years: ML 0.65–0.75 vs aMAP 0.73, 0.70, 0.68; no advantage in C-index, Brier score or decision curves |
| PRE-Screen (18) | general population, >900,000 individuals, 983 HCC; UK Biobank (development), All of Us (external test) | random forest on routine data, post-hoc explanations | no | published HCC risk scores | outperformed the scores; population prescreening, not surveillance in cirrhosis |
| **This study** | 17,177 adults with cirrhosis, all etiologies, region-wide routine EHR | three-class EBM, no interactions: the shape functions are the model | yes: a class at training; Blanche AUC($t$), Wolbers C, Aalen–Johansen, Fine–Gray at evaluation | aMAP, THRI at their own landmarks; binary EBM | $\Delta$AUC(12 mo) +0.10 / +0.06; washout $\Delta$C +0.06 / +0.04; fewer deaths per HCC flagged |

Supplementary Table 14. **Safeguards against real-world-data pitfalls,** mapped to the PROBAST+AI domains (participants, predictors, outcome, analysis) (19) and the TRIPOD+AI items (20).

| Pitfall | Why it biases | Our design | Evidence |
|---|---|---|---|
| Selection on future information | requiring follow-up or later records selects survivors | eligibility judged at time zero only (prediction date or landmark); one source population and one rule for development and evaluation | Methods; Fig. 1b |
| Misaligned time zero, immortal time | risk counted from a time patients had to survive to | follow-up starts at time zero (landmark or visit); each score's landmark is the first date it can be computed | Fig. 1a |
| Outcome-anchored sampling | case-control-like index dates misstate the case mix | evaluation at routine landmarks, every patient predicted by a model that never saw them, no retraining | Fig. 2; Table 3 |
| Leakage and protopathic bias | tests ordered on suspicion of HCC predict it | features end at time zero; 6-month washout | Table 3 washout columns; Supplementary Fig. 3 |
| Competing risks ignored | treating death as censoring overstates risk; as negative, it distorts risk factors | death or transplant is a class; Aalen–Johansen, Blanche AUC($t$), Wolbers C, Fine–Gray (21) | Figs 2–4; Supplementary Fig. 7 |
| Informative presence of tests | ordering carries information (22) | labs median-imputed, so a missing test contributes as a median value; testing intensity not a feature; washout; comparison by whether the key tests were measured | Methods; Supplementary Table 3 |
| Optimistic CV comparisons | fold dependence inflates significance | same 100 folds for every model; Nadeau–Bengio corrected t-test; Bayesian ROPE; Bonferroni ×3 | Table 2 |
| Uncalibrated, class-weighted probabilities | weights shift the intercepts (eq. 3) | Fine–Gray recalibration at lab visits, cross-fitted; Brier, IPA, O/E, slope; checked by sex and at time zeros that are not visits | Fig. 4d; Supplementary Fig. 5; Supplementary Tables 11–12 |
| Unfair comparisons with scores | different patients, times or recalibration | each score at its own landmark with all components measured; same patients; same recalibration; THRI bounded for unrecorded HCV cure | Fig. 2; Supplementary Fig. 2 |
| Post-hoc explanations | SHAP approximates a black box | glass box: the shape functions are the model; contrasts identified; deployed scorer returns exact contributions | Fig. 3; Fig. 4a–c |
| Subgroup performance | average gains hide groups left out | sex, age, etiology, prevalent or incident cirrhosis, key tests measured, landmark year, including where the gain vanishes (women, MASLD, missing tests) | Supplementary Fig. 3; Supplementary Table 3 |
| One region, one period | a model may learn local practice | leave-one-province-out refits, pooled by random effects; landmark year | Supplementary Tables 3 and 10 |
| Small-cell disclosure | re-identification | counts under 10 masked; only aggregates leave the secure server | all items |

69 # References


70 1. VAN HOUWELINGEN HC. Dynamic Prediction by Landmarking in Event History Analysis. Scandinavian Journal of Statistics. 2006;34(1):70–85. doi:10.1111/j.1467-9469.2006.00529.x

71 2. Blanche P, Dartigues J, Jacqmin-Gadda H. Estimating and comparing time-dependent areas under receiver operating characteristic curves for censored event times with competing risks. Statistics in Medicine. 2013;32(30):5381–97. doi:10.1002/sim.5958

72 3. Wolbers M, Blanche P, Koller MT, Witteman JCM, Gerds TA. Concordance for prognostic models with competing risks. Biostatistics. 2014 Feb;15(3):526–39. doi:10.1093/biostatistics/kxt059

73 4. Aalen OO, Johansen S. An empirical transition matrix for non-homogeneous Markov chains based on censored observations. Scandinavian Journal of Statistics [Internet]. 1978;5(3):141–50. Available from: https://www.jstor.org/stable/4615704

74 5. Kattan MW, Gerds TA. The index of prediction accuracy: an intuitive measure useful for evaluating risk prediction models. Diagnostic and Prognostic Research. 2018 May;2(1). doi:10.1186/s41512-018-0029-2

75 6. Fine JP, Gray RJ. A Proportional Hazards Model for the Subdistribution of a Competing Risk. Journal of the American Statistical Association. 1999;94(446):496–509. doi:10.1080/01621459.1999.10474144

76 7. Nadeau C, Bengio Y. Inference for the Generalization Error. Advances in Neural Information Processing Systems. 1999;12.

77 8. Benavoli A, Corani G, Demšar J, Zaffalon M. Time for a Change: a Tutorial for Comparing Multiple Classifiers Through Bayesian Analysis. Journal of Machine Learning Research [Internet]. 2017;18(77):1–36. Available from: http://jmlr.org/papers/v18/16-305.html

78 9. Fan R, Papatheodoridis G, Sun J, Innes H, Toyoda H, Xie Q, et al. aMAP risk score predicts hepatocellular carcinoma development in patients with chronic hepatitis. Journal of Hepatology. 2020 Dec;73(6):1368–78. doi:10.1016/j.jhep.2020.07.025 PubMed PMID: 32707225.

79 10. Sharma SA, Kowgier M, Hansen BE, Brouwer WP, Maan R, Wong D, et al. Toronto HCC risk index: A validated scoring system to predict 10-year risk of HCC in patients with cirrhosis. Journal of Hepatology. 2018 Jan;68(1):92–9. doi:10.1016/j.jhep.2017.07.033

80 11. Kanwal F, Lopez C, Ning J, Luster M, Reddy KR, Parikh ND, et al. Validation of Texas Hepatocellular Carcinoma Consortium Risk Index in the Hepatocellular Carcinoma Early Detection Strategy Study. Clinical Gastroenterology and Hepatology. 2026 Oct;24(10):2724–31. doi:10.1016/j.cgh.2026.01.041

81 12. Audureau E, Carrat F, Layese R, Cagnot C, Asselah T, Guyader D, et al. Personalized surveillance for hepatocellular carcinoma in cirrhosis – using machine learning adapted to HCV status. Journal of Hepatology. 2020 Dec;73(6):1434–45. doi:10.1016/j.jhep.2020.05.052

82 13. Ioannou GN, Tang W, Beste LA, Tincopa MA, Su GL, Van T, et al. Assessment of a Deep Learning Model to Predict Hepatocellular Carcinoma in Patients With Hepatitis C Cirrhosis. JAMA network open. 2020 Sep;3(9). doi:10.1001/JAMANETWORKOPEN.2020.15626 PubMed PMID: 32870314.

83 14. Kim HY, Lampertico P, Nam JY, Lee HC, Kim SU, Sinn DH, et al. An artificial intelligence model to predict hepatocellular carcinoma risk in Korean and Caucasian patients with chronic hepatitis B. Journal of Hepatology. 2022 Feb;76(2):311–8. doi:10.1016/j.jhep.2021.09.025

84 15. Nam JY, Sinn DH, Bae J, Jang ES, Kim JW, Jeong SH. Deep learning model for prediction of hepatocellular carcinoma in patients with HBV-related cirrhosis on antiviral therapy. JHEP Reports. 2020 Dec;2(6):100175. doi:10.1016/j.jhepr.2020.100175

85 16. Lin H, Li G, Delamarre A, Ahn SH, Zhang X, Kim BK, et al. A Liver Stiffness–Based Etiology-Independent Machine Learning Algorithm to Predict Hepatocellular Carcinoma. Clinical Gastroenterology and Hepatology. 2024 Mar;22(3):602–610.e7. doi:10.1016/j.cgh.2023.11.005

86 17. Nahon P, Layese R, Natella PA, Parlati L, Saidi T, Ganne-Carrié N, et al. Comparative evaluation of regression and machine-learning models for hepatocellular carcinoma risk stratification across diverse aetiologies. JHEP Reports. 2026 Apr;8(4):101740. doi:10.1016/j.jhepr.2026.101740

87 18. Clusmann J, Koop PH, Zhang DY, Haag F van, Nahhas OSME, Seibel T, et al. Machine Learning Predicts Hepatocellular Carcinoma Risk from Routine Clinical Data: A Large Population-Based Multicentric Study. Cancer discovery. 2026 May;16(7):OF1–19. doi:10.1158/2159-8290.CD-25-1323 PubMed PMID: 41881847.

88 19. Moons KGM, Damen JAA, Kaul T, Hooft L, Andaur Navarro C, Dhiman P, et al. PROBAST+AI: an updated quality, risk of bias, and applicability assessment tool for prediction models using regression or artificial intelligence methods. BMJ. 2025 Mar;388:e082505. doi:10.1136/bmj-2024-082505

89 20. Collins GS, Moons KGM, Dhiman P, Riley RD, Beam AL, Van Calster B, et al. TRIPOD+AI statement: updated guidance for reporting clinical prediction models that use regression or machine learning methods. BMJ. 2024 Apr;385:e078378. doi:10.1136/bmj-2023-078378

90 21. Geloven N van, Giardiello D, Bonneville EF, Teece L, Ramspek CL, Smeden M van, et al. Validation of prediction models in the presence of competing risks: a guide through modern methods. BMJ. 2022 May;e069249. doi:10.1136/bmj-2021-069249

91 22. Agniel D, Kohane IS, Weber GM. Biases in electronic health record data due to processes within the healthcare system: retrospective observational study. BMJ. 2018 Apr;k1479. doi:10.1136/bmj.k1479
